# Mental health in Germany before, during and after the COVID-19 pandemic

**DOI:** 10.1101/2024.06.21.24309286

**Authors:** Alexander Patzina, Matthias Collischon, Rasmus Hoffmann, Maksym Obrizan

## Abstract

Based on nationally representative panel data (N person-years=40,020; N persons=18,704; Panel Labour Market and Social Security; PASS) from 2018 to 2022, we investigate how mental health changed during and after the COVID-19 pandemic. We employ time-distributed fixed effects regressions to show that mental health (Mental Health Component Summary Score of the SF-12) decreased from the first COVID-19 wave in 2020 onward, leading to the most pronounced mental health decreases during the Delta wave, which began in August 2021. In the summer of 2022, mental health had not returned to baseline levels. An analysis of the subdomains of the mental health measure indicates that long-term negative mental health changes are mainly driven by declines in psychological well-being and calmness. Furthermore, our results indicate no clear patterns of heterogeneity between age groups, sex, income, education, migrant status, childcare responsibilities or pre-COVID-19 health status. Thus, the COVID-19 pandemic appears to have had a uniform effect on mental health in the German adult population and did not lead to a widening of health inequalities in the long run.

## 1 Introduction

Numerous studies indicate that the COVID-19 pandemic affected health outcomes in various ways that are heterogeneous between subpopulations of society (e.g., Blendermann et al. 2023; Cénat et al. 2021, 2022; Sun et al. 2023). Although the body of literature is vast, we identify three main issues in this literature. First, the majority of research focuses on the immediate or medium-term effects of the pandemic and leaves open the central question of whether health outcomes returned to prepandemic baseline levels. Second, much of the research on COVID-19 relies on convenience samples established during the COVID-19 pandemic. This constitutes a problem when investigating the longer-term effects of the pandemic on population health because in many cases, this sampling design does not allow for generalizable conclusions (e.g., Etikan et al. 2016). Third, empirical estimates of the heterogeneous effects of the COVID-19 pandemic stem from multiple research sources that rely on different samples and estimation methods, and the literature lacks consensus on the mode of analysis. Consequently, it is not entirely clear whether health inequalities widened, remained the same or even decreased during and after the COVID-19 pandemic. To improve our understanding of these heterogeneities, recent research emphasizes the need to provide evidence on heterogeneity in COVID-19 effects from one data source while holding the sampling and estimation design constant (Altmeijd et al. 2023).

This study addresses these research gaps and makes three central contributions. First, this study investigates the long-term effect of the COVID-19 pandemic on mental health. In particular, this study examines mental health changes in the prepandemic period, different phases of the pandemic between early 2020 and spring 2022, and summer 2022 (i.e., postpandemic). In doing so, we rely on a well-established health index that measures individuals’ mental health based on the Mental Health Component Summary Score from the SF-12 (Ware, Kosinksi, and Keller 1996). Thus, unlike most previous studies on COVID-19, we use a comprehensive index that covers four dimensions of mental and emotional well-being instead of focusing on a single dimension (e.g., depression). As prior research has demonstrated that mental health problems lead to decreased productivity (e.g., de Oliveira et al. 2023) and that good health leads to higher wages in the labor market (e.g., Jäckle and Himmler 2010), the focus of our study on the post-pandemic period also increases our understanding of unintended non-medical longer-term effects of the pandemic.

Second, we draw on a nationally representative panel study (Panel Labour Market and Social Security; see: Trappmann et al. 2019) with a large sample size and a long observation period. Based on this data set, we leverage within-person changes in mental health across different time points, which yields causal estimates under the assumption that sorting by survey interview dates (i.e., sorting into broader defined pandemic and post-pandemic periods) is exogenous. If this assumption holds, we identify the total effect of the pandemic, including for example lockdowns and fear of infection. The use of this design and data on mental health constitutes a significant contribution to the existing COVID-19 literature.

Third, in addition to investigating the overall effect of the COVID-19 pandemic on mental health changes, our study contributes to existing research that investigates heterogeneities by subgroups. We focus on dimensions that prior research identified as potential sources of heterogeneity. To this end, we investigate heterogeneity along the dimensions of sex, age, household income, migration background, education, childcare responsibilities and pre-COVID-19 health status (e.g., Almeida et al. 2020; Carrillo-Vega et al. 2020; Cénat et al. 2021; Connor et al. 2020; Elsayed et al. 2022, Entringer et al. 2020; Geng et al. 2021; Gibson et al. 2021; Möhring, Reifenscheid, and Weiland 2021; Patel et al. 2022; Xiong et al. 2020; Zoch, Bächmann, and Vicari 2021). In focusing on these heterogeneities, we advance the current state of research on potential health inequalities induced by the COVID-19 pandemic.

While the most obvious health effect of the COVID-19 pandemic is that individuals who contracted the disease suffered mild to severe symptoms (or died), focusing on mental health is important with regard to the societal dimension of the health crisis. In particular, people who were not infected with the coronavirus might also experience effects on different dimensions of their health. Counterintuitively, health may have even improved during the pandemic because individuals with poor health before the pandemic experienced a subjective relative improvement in health status, partly due to social comparison with infected individuals (Van de Weijer et al. 2022). In contrast, the health status of uninfected individuals might have deteriorated because of fear of contracting the disease, sorrow about friends and family members who died or became sick and general negative consequences of the pandemic. While it is possible that these factors also (indirectly) affect physical health, their main effect is on mental health through perceived risks, fear and stress (Wilson et al. 2020). Therefore, mental health is a valid and important dimension of the impact of COVID-19 on the health of the general population.

Our main results indicate that in the summer of 2022, mental health had still not entirely returned to prepandemic levels. The most severe health changes occurred during the Delta wave in 2021, which constituted the most dangerous phase of the pandemic in terms of pathogenicity (i.e., severity of illness) due to COVID-19 infections (e.g., Markov et al. 2023). Interestingly, our data do not indicate much heterogeneity across subgroups of the German population. Instead, the data indicate that individuals with good physical health prior to the pandemic had the most negative mental health changes in the first part of the pandemic (i.e., until summer 2021). Furthermore, our data suggest potential heterogeneity by age, with younger individuals appearing to be more affected during certain pandemic waves but not in the long run (i.e., health adaption is similar between younger and older individuals). Moreover, our data suggest potential heterogeneity between natives and migrants in 2020 and during the Delta wave. However, many of the heterogeneities we identify are not statistically significant from each other, potentially due to limited statistical power. Thus, while some heterogeneities existed in mental health changes during the pandemic, the pandemic appeared to have a uniform effect on mental health in the German population.

## 2 Previous research on COVID-19 and mental health

### 2.1 COVID-19 and mental health in Germany

According to Banks, Fancourt and Xu (2021), negative changes in mental health during the COVID-19 pandemic could be attributed to four main causes. First, individuals experienced health-related anxieties, such as the risk of being infected or hospitalized, which may have differed by an individual’s exposure and attitudes toward health risk. Second, there were financial concerns in the short and long run. Third, domestic living arrangements during the lockdown were sometimes a source of stress. Fourth, individual lifestyles were affected by the loss of social contacts and the transition to online social connections.

Evidence from systematic literature reviews unambiguously supports the claim that the COVID-19 pandemic has had a negative impact on many mental health domains, such as anxiety, depression, post-traumatic stress disorder, and psychological distress, worldwide (e.g., Salari et al 2020; Xiong et al. 2020). Important drivers of the adverse mental health impact appear to be experiences of isolation and quarantine that lasted longer than one week (e.g., Henssler et al. 2021).

Cross-sectional studies in Germany suggest that between March and July 2020, depression and anxiety risks, distress and psychological burdens increased (e.g., Ahrens et al. 2021; Bäuerle et al. 2020; Hetkamp et al. 2020; Petzold et al. 2020, Skoda et al. 2021). According to Benke et al. (2020), who conducted a cross-sectional study among 4,335 adults from Germany in April and May 2020 found that stringent restrictions due to lockdown measures, a substantial reduction in social contacts and pronounced perceived changes in life were associated with greater mental health impairment. In addition, Mata et al. (2021), who employed a sample of approximately 3,500 randomly selected participants representative of the German population, showed that more screen time, more snacking, and less physical activity were related to higher symptoms of anxiety, depression, and loneliness.

Smaller-scale studies with a longer time horizon provide additional insights for Germany. Based on an online survey of 1,903 respondents, Liu et al. (2021) showed that the average prevalence of psychological distress associated with the COVID-19 pandemic rose significantly from 24% in April (COVID-10 peak of the first wave) to 66% in September 2020 (first off-peak transmission period). Reis et al. (2023) used a survey of 2,203 respondents in Germany starting from March 2020 to show that the level of anxiety decreased while depressive symptoms increased. Elsayed et al. (2022) conducted an online questionnaire of 474 respondents from February to April 2021 in healthcare and community settings in the Ulm region of Germany and showed that 80.4% of participants had high levels of psychological distress.

In addition to the cross-sectional evidence, which solely draws on information during the COVID-19 pandemic, some studies for Germany exist using longitudinal data, which also includes prepandemic information on mental health. Based on the Socio-Economic Panel Study (SOEP), Entringer et al. (2020) showed that at the beginning of the COVID-19 pandemic in April 2020, symptoms of depression and anxiety had increased relative to 2019. However, levels in 2020 were comparable to mental health levels in 2016. Dragano et al. (2022) analyzed data from the German National Cohort Study (NAKO) for 161,849 participants who answered questions about their mental health between May and November 2020. The comparison with prepandemic health (data collected between 2014 and 2019) showed a 2.4% and 1.5% increase in the prevalence of moderate or severe symptoms of depression and anxiety, respectively. The authors identified labor market processes such as becoming unemployed or changes in employment security as important drivers of the increases in symptoms.

### 2.2 Previous findings on heterogeneous effects

The literature on the heterogeneous effects of COVID-19 remains mostly limited and inconclusive. In particular, previous studies suggest that the COVID-19 pandemic increased inequality in mental health along the dimensions of income, education, ethnicity, migrant and minority status, and social isolation (e.g., Gibson et al. 2021, Parenteau et al. 2022, Patel et al. 2022). However, there are only a handful of studies on each dimension of inequality, and they often present conflicting evidence. For example, some studies indicate that individuals with higher levels of education and income showed better coping (Elsayed et al. 2022), while others claim that individuals with high education and high income reported a slight decrease in life satisfaction compared to those with low education and low income, demonstrating a slight increase in life satisfaction (Entringer et al. 2020).

The most systematic inequalities have been identified for sex, age and preexisting conditions in mental health impairments, which we discuss further in this subsection. According to previous systematic reviews, mental health for individuals under age 40 and women decreased in the first phase of the COVID-19 pandemic (e.g., Almeida et al. 2020; Xiong et al. 2020; Cénat et al. 2021). In particular, research indicates an increasing prevalence of generalized anxiety disorder, depressive symptoms, post-traumatic stress symptoms and poor sleep quality during the COVID-19 pandemic among young individuals and women (e.g., Huang and Zhao 2020; Jacques-Avino et al 2020; Rossi et al. 2020). In a more recent review of studies involving 50,000 or more participants, Penninx et al. (2022) report that the effect of the pandemic was heterogeneous and showed a statistically significant but small increase in self-reported mental health problems. Sun et al. (2023) provide evidence from a systematic review and meta-analysis of 134 cohorts collected prior to April of 2022 that no changes were found in general mental health, but symptoms of depression worsened minimally.

Additionally, findings from the UK indicated sizeable increases in scores on the 12-item General Health Questionnaire distress measure (GHQ-12), which were pronounced among females and the younger population (Pierce et al. 2020). Further longitudinal evidence from the UK comparing prepandemic levels of mental health to levels in the first month of the UK lockdown in 2020 corroborated these findings (Proto and Quintana-Domeque 2021). Evidence from Wales covering a later period also indicated greater effects of the COVID-19 pandemic on psychological distress among females and the younger population (Gray et al. 2020). Research on the United States, which used three waves of geographically representative survey data in March, April and May 2020, indicated that the negative mental health effect of lockdown measures was entirely driven by women and cannot be explained by an increase in financial worries or caregiving responsibilities (Adams-Prassl et al. 2020a). In addition to these rather short-term studies, early research also addressed longer-term consequences of the COVID-19 pandemic on the mental health of women and young adults (e.g., Henseke, Green, and Schoon 2022; Blendermann et al. 2023; Sandner et al. 2023; Zwar, König, and Hajek 2023). However, evidence of sex and age inequality based on nationally representative panel data, pre–post comparisons, and a longer study period encompassing the second year of the pandemic remains limited.

At the beginning of the COVID-19 pandemic, research indicated that individuals with a history of mental health problems or individuals with chronic illness showed lower levels of mental health (McCracken et al. 2020; Xiong et al. 2020). More recent studies also indicate that individuals with preexisting conditions (i.e., chronic illnesses or poor health in 2019) were at risk for poor mental health during the second wave of the COVID-19 pandemic that started at the end of 2020 (Buneviciene et al. 2022). Another recent study focusing on young adults and analyzing retrospective data covering the time before and during the pandemic from February 2022 (i.e., individuals ages 18 to 21) indicates that the anxiety and depression risks of individuals without preexisting mental health problems increased over the course of the pandemic (Kleine et al. 2023). Blendermann et al. (2023) report in their systematic review that pre-existing mental health diagnoses were not associated with symptom exacerbation, except for obsessive-compulsive disorder. Thus, it remains an empirically open question whether poor health before the COVID-19 pandemic buffers or amplifies the impact of the pandemic on mental health.

## 3 Methods

### 3.1 Data

This study uses data from the Panel Labour Market and Social Security (PASS; see: Trappmann et al. 2019). The panel has been surveying approximately 10,000 German households and approximately 15,000 individuals since 2006. The PASS is a representative general population sample. In our analysis, we restrict the PASS sample to the years 2018 (field time: 14.02.2018 to 15.09.2018), 2020 (field time: 14.02.2020 to 27.09.2020), 2021 (field time: 11.02.2021 to 19.09.2021), and 2022 (field time: 16.02.2022 to 13.09.2022). As our outcome measure was not part of the survey in 2019, a substantial part of pre-COVID-19 health information stems from 2018. The PASS data are ideally suited for investigating mental health changes during the COVID-19 pandemic because due to its panel structure, the PASS includes pre-COVID measures. Thus, the dataset enables us to compare responses before and after the outbreak of the pandemic. Our analytical strategy will exploit this key feature of the data.

### 3.2 Measures

#### Mental health

To measure mental health, we used a well-established index variable measuring the mental health of individuals based on the Mental Health Component Summary Score from the SF-12 (Ware, Kosinksi, and Keller 1996). This index ranges from 1 (poor) to 5 (very good) and measures mental health for the four weeks prior to the survey. The survey questions in the PASS to retrieve this index operationalize vitality, role restrictions due to emotional problems (which we label emotional stability and is often labeled ‘role emotional’; see Ware, Kosinksi, and Keller 1996), psychological distress and wellbeing (Ware, Kosinksi, and Keller 1996). Note that the domain of social functioning is missing in the PASS data. However, the latent constructs of vitality and social functioning are correlated, so the constructed index should still approximate individuals’ overall mental health. The information used stems from answers to the following questions: “Please recall the past four weeks. How often did it happen during this time that… (a) you felt melancholic and depressed?, (b) you felt calm and balanced?, (c) you felt highly energetic?, (d) that you did less at work or in your daily life than you intended to because of mental or emotional problems?” The items show a high internal validity (Cronbach’s Alpha=0.77). After reverse scoring items (a) and (d), we created a sum score for each individual in the dataset. As the mental health measure captures cognitive and affective parts of mental health, this outcome constitutes a valid measure of individuals’ mental health.

#### Time (pandemic phases)

Because we are interested in how health outcomes changed during the pandemic, the main independent variable for our analysis is time. Instead of simply using a running time axis, we cluster the used time variable to describe changes in health according to pandemic phases. Information for this categorization of time comes from the Robert Koch Institute in Germany (RKI 2022). An overview of the clustering and the distribution in the dataset is presented in Table 1.

**Table 1.** Overview of employed time (explanatory) variable

| **Pandemic Phase** | **Calendar Weeks/Months/Year** | **Share of Observations in the Sample** |
| --- | --- | --- |
| **Before** | 7 to 37/February to September/2018 | 32% |
| **First wave** | 6 to 20/February to May/2020 | 4% |
| **Summer 2020** | 21 to 39/May to September/2020 | 11% |
| **Second wave** | 40 to 8/September to February/2020-2021 | 3% |
| **Alpha wave** | 9 to 23/February to June/2021 | 21% |
| **Summer 2021** | 24 to 30/June to July/2021 | 2% |
| **Delta wave** | 31 to 51/August to December/2021 | 2% |
| **Omicron Wave** | 52 to 21/December 2021 to May 2022 | 21% |
| **Summer 2022** | 21 onwards/May 2022 onwards | 5% |

*Source:* Own presentation based on RKI (2022).

The baseline value “before” uses information from 2018. Thus, baseline measures of mental health are not affected by the COVID-19 pandemic. The second period of our time variable includes calendar weeks 6 to 20 in 2020 and corresponds with the “first wave”, i.e., the initial COVID-19 shock that comprises the first nationwide lockdown between calendar weeks 13 and 18 (22.03.20-03.05.20). The next pandemic phase comprises calendar weeks 21 to 39, which we label “summer of 2020”. This period represents a time with a low incidence of COVID-19 cases and a time of relaxation within German society. The next phase of the pandemic comprises the “second wave” and represents the period between calendar weeks 40 in 2020 and 8 in 2021. During the second COVID wave, a new subvariant (i.e., B.1.1.7; see RKI 2021) of the virus emerged, and virologists and epidemiologists identified a new COVID wave. Consequently, the next pandemic phase represents the “Alpha wave” and covers the period between calendar weeks 9 and 23 in 2021. The “second” and “Alpha” waves represent the first hard COVID winter and spring in Germany. During these waves, the German government implemented a partial lockdown between calendar weeks 45 and 51 (02.11.20-15.12.20) and a very strict lockdown between calendar weeks 53 and 9 (28.12.20-03.03.21). Afterward (between calendar weeks 10 and 15; 08.03.21-18.04.21), German federal states started to relax precautionary measures again.

The next value of our pandemic phase indicator includes the summer of 2021 (calendar weeks 24 to 30), which again represents an era of relaxation within German society. The next period our data cover is the Delta wave, which started in calendar week 31 in 2021 and lasted until the end of 2021. Subsequently, the Omicron variant became prevalent in Germany. The Omicron wave lasted until calendar week 21 in 2022. We label the last observed period in our data, the time from May 2022 onwards, as Summer 2022. This constitutes the first post-pandemic period in Germany because hospitalization rates declined rapidly at the beginning of spring 2022, and the virus started to become endemic. Note, however, that some legal restrictions (e.g., mask wearing in trains) were still in place.

#### Moderators

An additional aim of this research was to explore heterogeneity in the change in mental health during the COVID-19 pandemic between population subgroups. To this end, we used the survey answers of respondents to categorize individuals by sex. Additionally, prior research indicated that adolescents and young adults were strongly hit by the pandemic (e.g., Henseke, Green, and Schoon 2022; Sandner et al. 2023). Therefore, we categorized individuals into two age groups, below age 35 and 35 or older. Moreover, we explored heterogeneity in mental health changes according to pre-COVID physical health measured in 2018. We categorized individuals as healthy if they indicated very good or good health in the standard and widely used self-rated health item (Mossey and Shapiro 1982). Furthermore, we investigate heterogeneity by education (tertiary education vs. lower), income (below and equal or above median equivalent household income in 2020; the median is 1333€); migration (native vs. any migration background) and children below the age of 15 in the household.

#### Control variables

At each analytical step, we include the survey mode and interview month as control variables. The survey mode is important because over the course of the pandemic, the PASS switched all in-person interviews to telephone (assisted) interviews. Additionally, interview month constitutes a crucial control variable because mental health measures suffer from seasonality (e.g., Zhang et al. 2021). Finally, age is a crucial control variable because mental health systematically changes with individuals’ age (see, for example, Bell 2014), which we include in each analytical step.

### 3.3 Sample

Our analytical sample (see Table 2) includes observations with valid information in all measures (We did not use a multiple imputation procedure because most missing information stemmed from the variable on health status in 2018, which is prone to panel attrition. Thus, we would have to impute information on the outcome for this subgroup, which is problematic (see for example, Dupre 2007)). Table 2 gives an overview of the distributions of all variables used in the analysis. This table already indicates that mean health declined over the course of the pandemic. Moreover, 50% of PASS interviews took place before mid-April of a calendar year before and during the COVID-19 pandemic. Additionally, approximately 58% of the interviews were CAPI interviews, while 42% were CATI interviews. Over the course of the pandemic, the share of CATI interviews increased, and CAPI interviews took place mainly via telephone. While the distribution of interview modes changed slightly during the pandemic, in the case of PASS, the pandemic did not induce a mode change, which provided survey participants more privacy. This is important because changes to more personal interview situations could heavily influence participants’ responses to sensitive questions such as the health outcomes under study.

**Table 2.** Distribution of outcome, moderating and control variables across analytical data sets

|  | Overall |  | Before<br>2020 |  | 2020<br>to<br>2022 |  |
| --- | --- | --- | --- | --- | --- | --- |
|  | Mean | SD | Mean | SD | Mean | SD |
| Mental health (Mental Component Summary 'MCS' of SF-12; 1 to 5) | 3.52 | 0.84 | 3.61 | 0.84 | 3.47 | 0.84 |
| *Subdomains of MCS* |  |  |  |  |  |  |
| Psychological well-being | 3.64 | 1.08 | 3.75 | 1.09 | 3.58 | 1.07 |
| Feeling calm | 3.41 | 1.06 | 3.46 | 1.07 | 3.38 | 1.05 |
| Vitality | 3.01 | 1.06 | 3.04 | 1.09 | 3.00 | 1.05 |
| Emotional stability | 4.01 | 1.11 | 4.21 | 1.07 | 3.92 | 1.11 |
| *Inequality domains* |  |  |  |  |  |  |
| Female (0/1) | 0.50 | - | 0.50 | - | 0.50 | - |
| Age (years) | 46.63 | 16.90 | 45.61 | 16.97 | 47.12 | 16.85 |
| Good pre-COVID-19 health status | 0.43 | - | 0.45 | - | 0.41 | - |
| Migrant (0/1) | 0.36 | - | 0.36 | - | 0.35 | - |
| High income (0/1) | 0.49 | - | 0.43 | - | 0.51 | - |
| Child under 15 (0/1) | 0.26 | - | 0.28 | - | 0.25 | - |
| High education | 0.21 | - | 0.20 | - | 0.22 | - |
| *Covariates* |  |  |  |  |  |  |
| Interview month (2-9) | 4.33 | - | 4.10 | - | 4.44 | - |
| Mode: CATI | 0.42 | - | 0.32 | - | 0.47 | - |
| Mode: CAPI, face to face | 0.24 | - | 0.68 | - | 0.03 | - |
| Mode: CAPI, by phone | 0.34 | - | 0.00 | - | 0.50 | - |
| Observations | 40020 |  | 12892 |  | 27128 |  |

Data: PASS data.

Table 2 also shows that the analytical sample is sex balanced, which did not change over the course of the COVID-19 pandemic. Additionally, the mean age is approximately 46 years, which is close to the German population mean. Moreover, 43% of respondents in the analytical sample reported a (very) good pre-COVID-19 health status. This share is more or less stable when comparing prepandemic with pandemic values.

### 3.4 Statistical analyses

In our empirical approach, we first show descriptive mean trends in our outcome variable over time. Second, we rely on individual fixed effects (FE) regressions to estimate intraindividual change over time (Allison 2009). This strategy has the key advantage that derived estimates only suffer from bias originating from time-varying unobserved heterogeneity, while other methods such as ordinary least square or random effects models rely on a stronger exogeneity assumption (i.e., no unobserved heterogeneity). By demeaning the data (i.e., only investigating deviations within individuals from individual-specific means), the FE estimates are not prone to bias originating from time-constant unobserved heterogeneity. However, the demeaning of the data comes at the cost of not being able to investigate levels and differences in levels between groups. To overcome this limitation, we opted to present both descriptive trends and estimates from the FE approach, thereby informing about level differences as well as how health was causally affected by COVID-19 and changed over the course of the pandemic. We estimate the following equation:

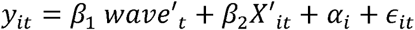

where *y_it_* is the outcome of interest, wave′*_t_* is a set of indicator variables for the specific pandemic phase, *X* is a set of covariates (indicators for interview months, interview mode and a continuous variable for age), *α_i_* is a set of individual-specific fixed effects and *∈_it_* is an idiosyncratic, time-varying error term. We argue that employing the FE approach to our setting delivers causal estimates of health changes. The main underlying assumption for this claim is that there is no systematic sorting into our employed time variable. As we categorized our time variable according to the progress of the pandemic, this variable is almost exogenous into certain interview dates occurs. As the chosen time windows in our time variable were rather broad, it is unlikely that sorting into interview dates heavily distorted estimates from the FE approach. Additionally, employing the FE estimator ensured that all time-constant potential selection variables into certain interview dates (e.g., sex, education, personality, etc.) were controlled for. Thus, under these assumptions, the FE analysis delivered causal effects of the pandemic on mental health between 2018 and the summer of 2022.

## 4 Results

### 4.1 Descriptive evidence: The development of mental health over time

Before showing results from fixed effects regressions, we descriptively investigate trends in mental health over time. To this end, Figure 1 shows trends in mental health beginning in 2018. In 2018 (i.e., before the COVID-19 pandemic), mean population mental health was approximately 3.61 scale points. Figure 2 indicates that during the first wave of the pandemic, mental health did not change on average, and some minor reductions in mental health occurred over the summer of 2020. During the second phase of the COVID-19 pandemic, i.e., during the second and Alpha waves, population mental health substantially decreased to approximately 3.42 scale points. During the summer of 2021, mental health recovered, but it declined again during the Delta wave, which began in August 2021. Subsequently, mental health almost returned to baseline levels in the summer of 2022, reaching 3.55 scale points.

**Figure 1.**
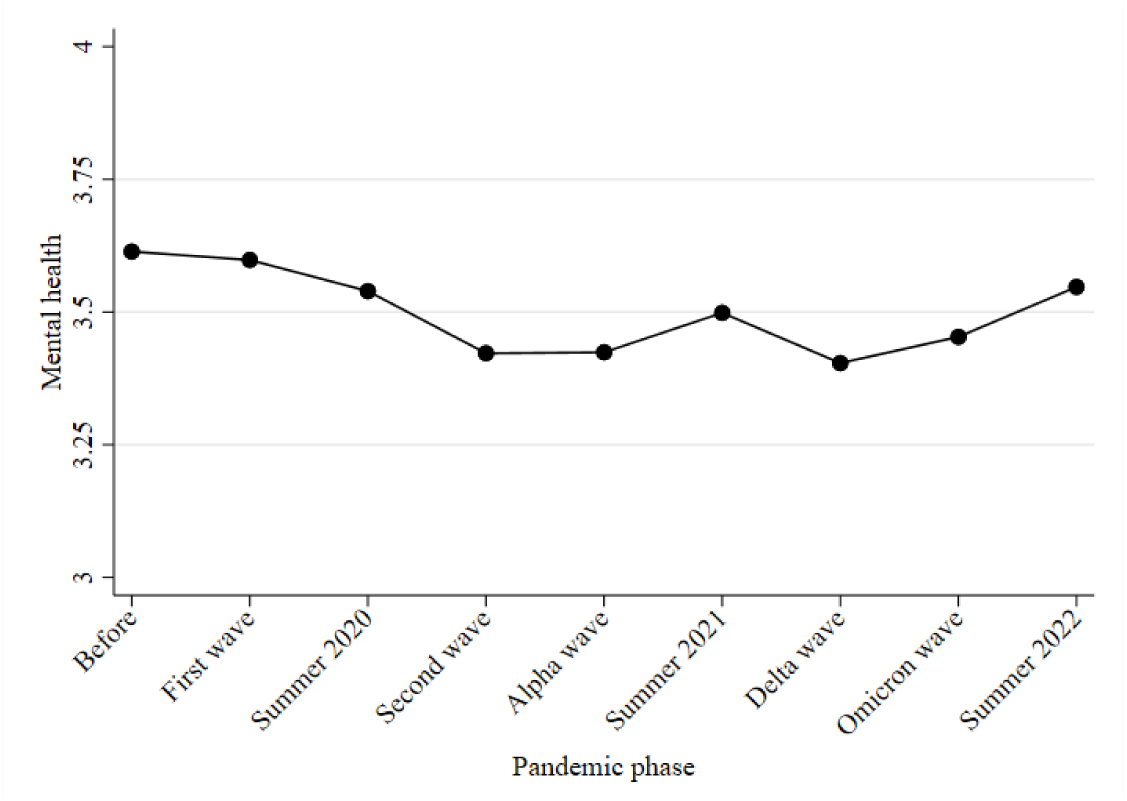
Mean development of mental health (on a scale from 1 to 5) before and during the COVID-19 pandemic

**Figure 2.**
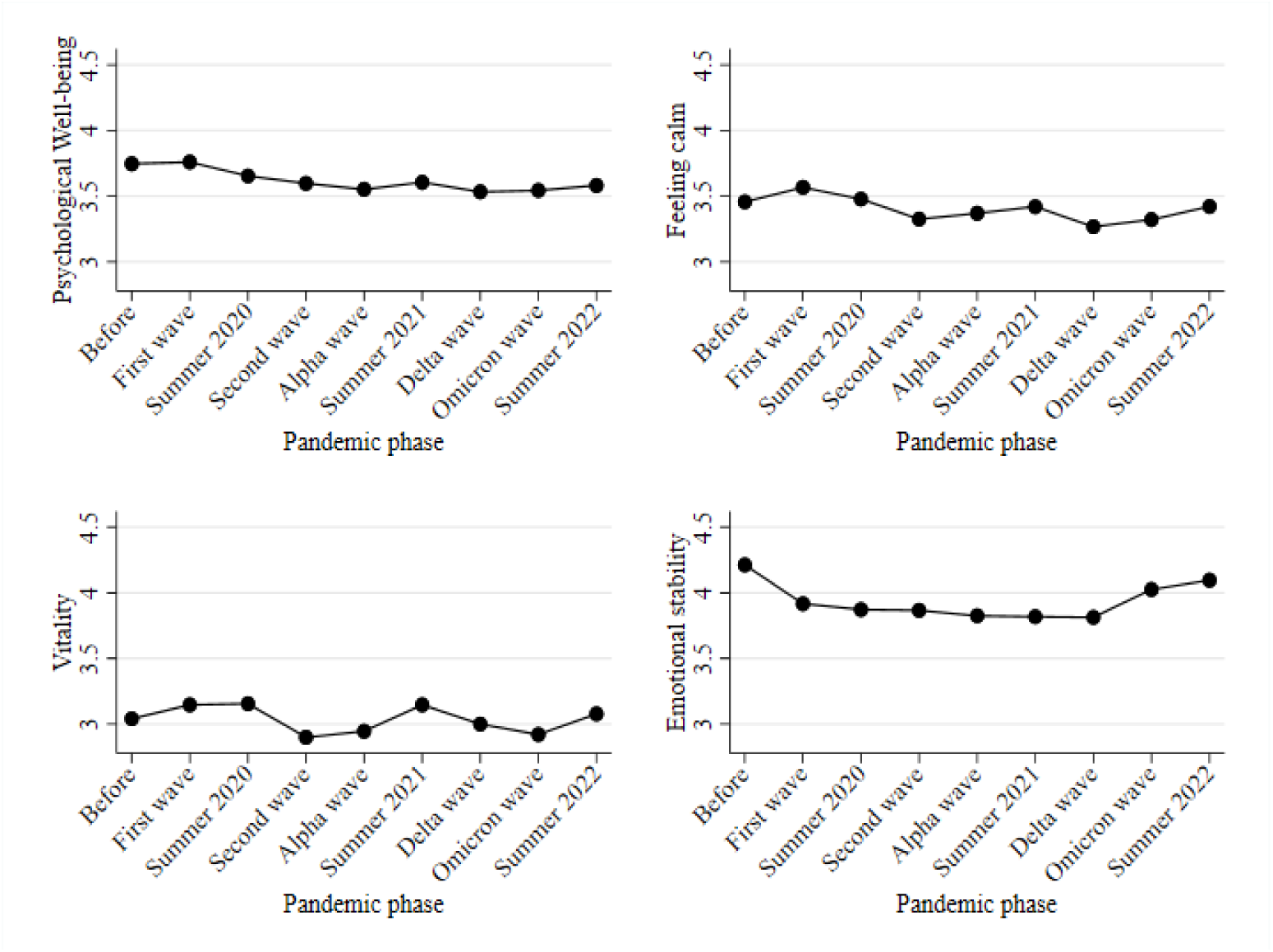
Mean development of mental health index components (on a scale from 1 to 5) before and during the COVID-19 pandemic

Next, in Figure 2, we investigate the development of the four mental health subdomains that are part of the overall mental health index. The upper-left part of Figure 2 indicates that psychological wellbeing (measured with melancholic and depressed feelings; here reversed so that higher values indicate higher levels of wellbeing) steadily decreased over the course of the pandemic. The trend during the pandemic mainly follows the development of the overall mental health index described in Figure 1. Interestingly, psychological well-being appears to remain at its prepandemic levels. The upper-right part of Figure 2 indicates that feelings of calm did not change substantially between the prepandemic period and summer of 2020. During the second wave, feelings of calm decreased on average, indicating increases in psychological distress. While feelings of calm slightly increased after the second wave, those feelings decreased again during the Delta wave. During the Omicron wave and the summer of 2022, feelings of calm returned to baseline levels.

The lower-left part of Figure 2 indicates that vitality slightly increased during the first wave of two COVID-19 waves in Germany and increased again during the summer of 2021. During the Delta wave, vitality levels decreased back to pre-COVID-19 levels and slightly increased again during the summer of 2022. The lower-right part of Figure 2 indicates a pronounced decrease in emotional stability (i.e., an increase in emotional problems) when comparing levels during the pandemic with levels before COVID-19. Additionally, our findings suggest that emotional stability particularly decreased during the first wave of the pandemic and only moderately decreased thereafter. During the Omicron wave and the summer of 2022, emotional stability returned almost to baseline levels.

### 4.2 Findings from fixed effects regressions: Intraindividual changes in mental health during pandemic phases

Figure 3 presents findings on how mental health changed during the COVID-19 pandemic in relation to before COVID-19 (see Table A.1 in the Appendix for regression tables). The upper-left part of Figure 3 shows overall changes in mental health during the COVID-19 pandemic. Our results indicate statistically significant negative mental health changes during the first wave and summer of 2020. During these periods, mental health decreased by 0.12 points during the first wave and 0.13 during the summer of 2020. These negative changes represent approximately 15% of a standard deviation (SD) in mental health. Thus, the initial changes were already not negligible. This is consistent with the systematic review and meta-analysis of longitudinal studies by Cénat et al. (2022), who reported that mental health problems peaked in April and May of 2020.

**Figure 3.**
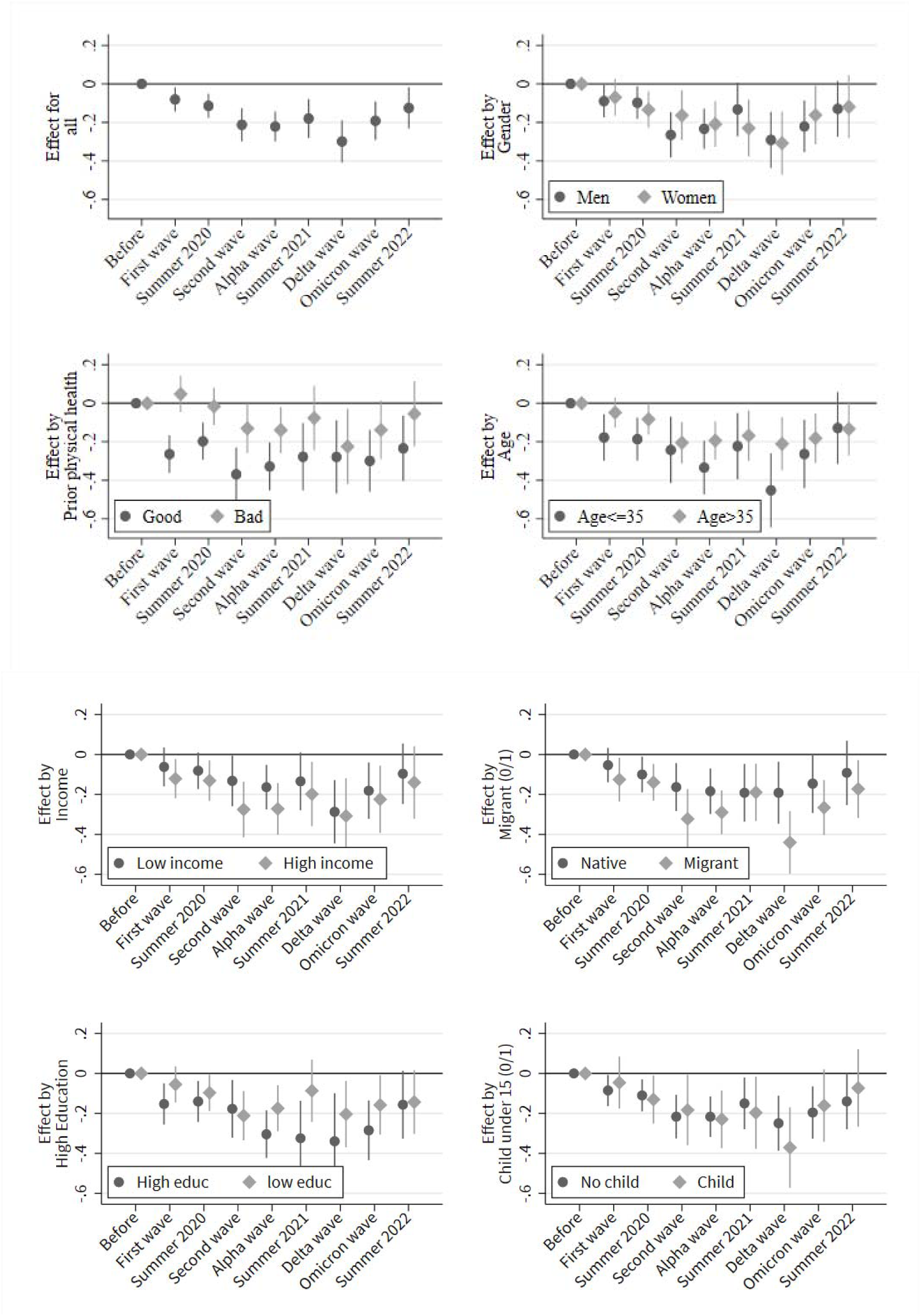
Heterogeneity in intra-individual changes of the mental health index during the pandemic. *Note.* Figure shows point estimates of the employed time variable based on individual fixed effects regressions. Control variables: Interview month indicator variables, interview mode, and age (linear).

During the second wave and until summer 2021, negative health changes intensified and amounted to approximately 27% of a SD in mental health. A further decrease in mental health occurred during the Delta wave, and negative mental health changes accounted for approximately 40% of a SD. Note that coefficients are less precisely estimated for the summer 2021 and Delta waves due to relatively small samples. During the Omicron wave, mental health improved again (approximately 23% of SD) but remained under baseline levels until summer 2022. In the first postpandemic period, negative health changes amounted to approximately 15% of a SD. We present the effect of the pandemic waves on other health outcomes in Figure A.1. The findings are ambiguous and show that the mental health scale of the SF-12 is distinct from health satisfaction (0-10), mental health problems (measured with a single-item question; 1-5) and self-rated health (1-5).

Parts of Figure 3 explore heterogeneous effects in the overall pattern. All parts revealed no statistically significant heterogeneity by sex, pre-COVID-19 health status, age, income, education, migration status and children. Note, however, that individuals with poor pre-COVID-19 physical health status do not show a significant mental health change in summer of 2021, while mental health statistically significantly decreases for individuals with good prepandemic health status. Additionally, our results suggest that during the Delta wave, the mental health of the younger population decreased more strongly than that of the older population (i.e., the decrease represents approximately 64% of a SD in mental health). However, this pronounced decrease for the younger population is not statistically significantly different from the change in the older population during the Delta wave. Moreover, our data suggest potential heterogeneity between natives and migrants in 2020 and during the Delta wave. In the long run, however, the COVID-19 pandemic had no effect on inequality in mental health as no differences across subgroups were found in the summer of 2022.

Figure 4 presents findings on how different components of mental health changed during and after the COVID-19 pandemic in relation to before COVID-19 (see Table A.2 in the Appendix for regression tables). While no substantial or statistically significant changes in vitality occurred (lower-left part of Figure 4), psychological well-being steadily decreased over the course of the pandemic. While changes in this mental health component were small during the first wave and summer of 2020, negative changes during the Delta wave were pronounced (approximately 0.31% of a SD in psychological well-being). Psychological well-being recovered slowly during the Omicron wave and remained under prepandemic levels in summer 2022 (approximately 14% of a SD). A similar pattern occurred in the domain ‘feeling calm and balanced’ (upper-right part of Figure 4). In the summer of 2022, feelings of calm were still below baseline levels (approximately 16% of a SD). The most pronounced changes during the pandemic occurred for emotional problems (lower-right part of Figure 4). Pronounced negative changes (approximately 34% of a SD) occurred at the beginning of the pandemic and further intensified during the Delta wave (approximately 42% of a SD). In contrast to feelings of calm and psychological well-being, emotional stability almost reached baseline levels during the Omicron wave and summer 2022. Appendix Figures A2 to A5 show the subgroup estimations for the domains.

**Figure 4.**
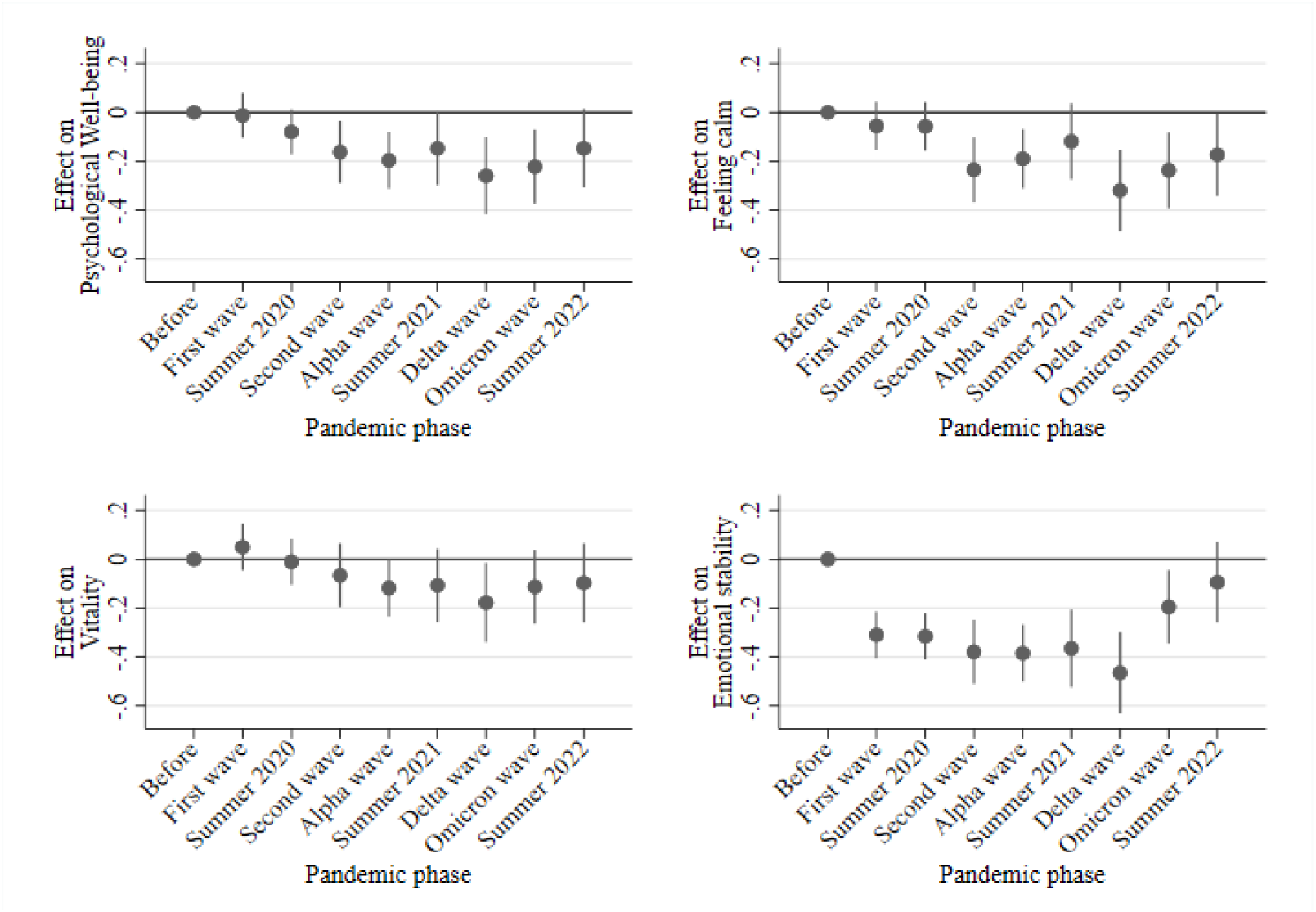
Intra-individual changes of the mental health components during the pandemic. *Note.* Figure shows point estimates of the employed time variable based on individual fixed effects regressions. Control variables: Interview month, interview mode, and age (linear).

## 5 Discussion & conclusion

Based on nationally representative longitudinal data from the Panel Labour Market and Social Security (PASS) and linear- and time-distributed fixed effects regressions, this study investigated whether mental health (measured with the MCS of the SF-12) changed during and after the COVID-19 pandemic in Germany. Specifically, this study investigated mental health changes from before the COVID-19 pandemic to different pandemic phases until the summer of 2022. The study expected to find negative mental health changes because the COVID-19 pandemic has had direct and indirect effects on mental health. Fears and sorrows may lead to negative health changes (e.g., Wilson et al. 2020). Additional indirect effects may emerge due to unintended effects of policies implemented to combat the pandemic (e.g., Aknin et al. 2022). Research on COVID-19 and mental health generally finds support for these theoretical mechanisms (e.g., Banks, Fancourt and Xu 2021). Our study contributes to this research and found the following results:

First, our descriptive results indicate a decline in mental health from before the COVID-19 pandemic that lasted until summer 2022. While the mental health average was approximately 3.6 scale points in 2018, it declined to 3.4 scale points in the Delta wave in Germany and recovered to 3.5 in summer 2022. The descriptive results show that population-averaged mental health started declining during the second phase of the COVID-19 pandemic and substantially decreased during the Delta wave. The descriptive results also indicate that psychological wellbeing (measured with melancholic and depressed feelings) steadily decreased over the course of the pandemic. Feelings of calm and balance did not change between 2018 (i.e., before the pandemic) and summer of 2020. However, during the second phase of the pandemic, such feelings decreased, indicating elevated levels of psychological distress. Furthermore, vitality remained rather stable during the pandemic. We found the most pronounced changes in levels of emotional stability. These changes were most pronounced during the first wave of the pandemic. Afterward, emotional stability decreased only slightly until the Delta wave in 2021. Interestingly, while emotional stability almost returned to prepandemic levels, psychological well-being remained at lower levels in summer 2022.

Second, our findings based on time-distributed fixed effects regressions corroborate the main findings from the descriptive workaround. Interestingly, and in contrast to the descriptive approach, findings from the fixed effects regressions indicated negative mental health changes during the first wave of the COVID-19 pandemic. From the first COVID-19 wave onward, mental health decreased, leading to the most pronounced changes during the Delta wave, which began in August 2021. These mental health changes appear to be mainly driven by increases in emotional problems. In the summer of 2022, mental health remained low and did not return to prepandemic levels. In contrast to much research on COVID-19 and mental health, our findings do not indicate a clear pattern of heterogeneity.

Overall, our findings indicate that the COVID-19 pandemic appears to have had a uniform effect on mental health changes in the German adult population, and inequalities did not increase depending on the severity during the pandemic (e.g., Maffly-Kipp et al. 2021) or in the longer run (i.e., in the first summer after the COVID-19 pandemic). We did not find pronounced heterogeneity among the adult population while relying on a large representative dataset, which implies that the effects of the COVID-19 pandemic on inequalities in mental health may be very complex and affect multiple subgroups of the population differently. Employing a representative dataset and a method that accounts for time-constant unobserved heterogeneity between subgroups might produce more valid results than earlier studies that may suffer from confounding bias. Moreover, research on COVID-19 and health has begun to investigate smaller subgroups of general populations. For instance, an elaborate research stream focusing on young individuals emerged, which unambiguously shows that adolescents have been severely hit by the pandemic (see, for example, Henseke, Green, and Schoon 2022; Magson et al. 2021; Neugebauer et al. 2024; Thorisdottir et al. 2021). Our results also suggest that during the Delta wave, the mental health of the younger population compared to the older population may have declined more strongly. Nevertheless, overall, our analysis suggests a rather uniform effect of the pandemic on mental health among adults in Germany.

Additionally, our study implies that analyses of the COVID-19 pandemic should rely on strong methodological designs. Our study showed that even if researchers are able to compare pre- and post-COVID-19 health, negative health changes may be disguised due to unobserved heterogeneity. Employing a fixed effects approach, which accounts for time-constant unobserved factors, revealed that mental health had already started to decrease during the first wave of the pandemic in Germany. In contrast, the simple mean comparison of pre-COVID-19 health and mental health during the first phase of the pandemic in Germany did not reveal such changes. Additionally, the employed fixed effects approach also revealed that the descriptive approach underestimated the negative health changes. These underestimations are particularly pronounced during the Delta wave in 2021. While the analysis of population-averaged mean changes revealed a decrease of approximately 23% of a SD, the fixed effect analysis indicated negative mental health changes were approximately 40% of a SD in mental health. Thus, the descriptive approach severely underestimated the negative consequences of the COVID-19 pandemic on mental health in Germany.

Although our study has many strengths, some weaknesses are apparent. In our analytical approach, we could not disentangle the direct and indirect effects of the pandemic. This is mainly due to data limitations because a detailed analysis of the impact of lockdowns, for example, would require a large number of cases to enable us to use temporal and regional variation for empirical identification. Moreover, direct measures of COVID-19 fear or worries are not available in the data, which would have enabled us to directly test our theoretical claims.

Additionally, we only focused on some dimensions of possible inequalities. For instance, we did not account for the effect of work arrangements (Hecker et al. 2020) or occupations (e.g., Dragano et al. 2022). Additionally, it is possible that we would have found inequalities if we had investigated mental health changes along more fine-grained lines, for example, by contrasting mental health changes for students with those of the working and retired populations. However, given our dataset, such an analysis was not possible. Future work must consider the possible intersectionality of inequalities (e.g., Bowleg 2020; Maestripieri 2021), which may be a cause of adverse mental health changes during (and after) the COVID-19 pandemic.

## Supporting information

Replication Syntax

## Data Availability

All data produced in the present study are available upon reasonable request to the Institute for Employment Research

https://fdz.iab.de/en/pd_hd/panel-study-labour-market-and-social-security-pass-version-0622-v1/

## Appendix

**Figure A1.**
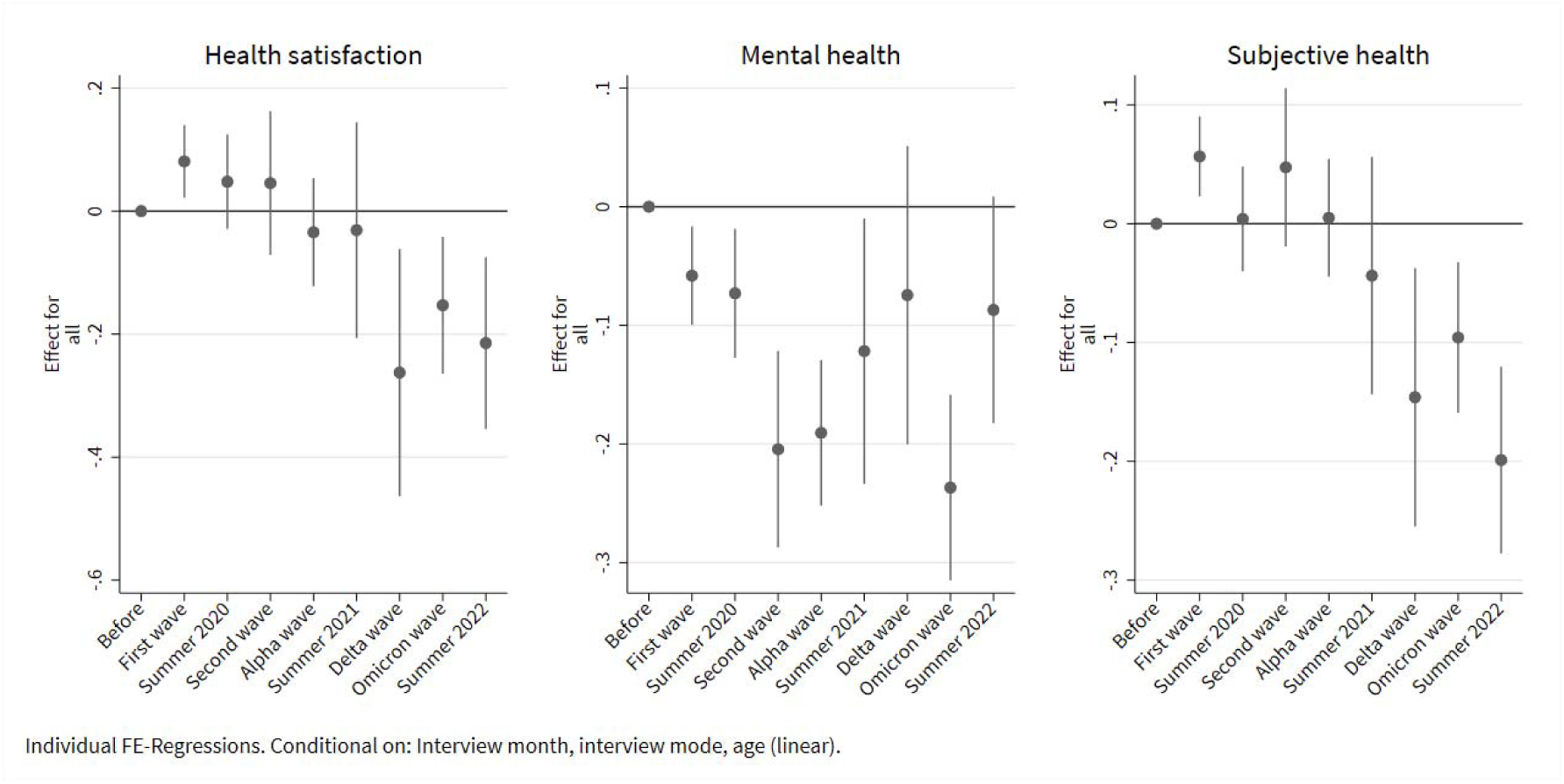
Effects on other outcomes. *Note:* Figure shows point estimates of the employed time variable based on individual fixed effects regressions. Control variables: Interview month, interview mode, and age (linear). The left-hand side of this figure shows the results of health satisfaction measured with a single-item question (“How satisfied are you today with the following areas of your life andHow satisfied are you with your health?”) ranging from 0 to 10. The middle part of this figure shows the results for mental health problems (“How strongly have you been affected by mental problems, such as fear, dejection or irritability in the past 4 weeks? Please tell me whether you were affected (1) ‘Extremely’; (2) ‘Quite a bit’; (3) ‘Moderately’; (4) ‘A little bit’; (5) ‘Not at all’”) ranging from 1 to 5. The right-hand side of this figure shows the results for self-rated health (“How would you describe your state of health in the past 4 weeks in general? Was it (1) ‘Bad’; (2) ‘Not so good’; (3) ‘Satisfactory’, (4) ‘Good’; (5) ‘Very good’”) ranging from 1 to 5.

**Figure A2.**
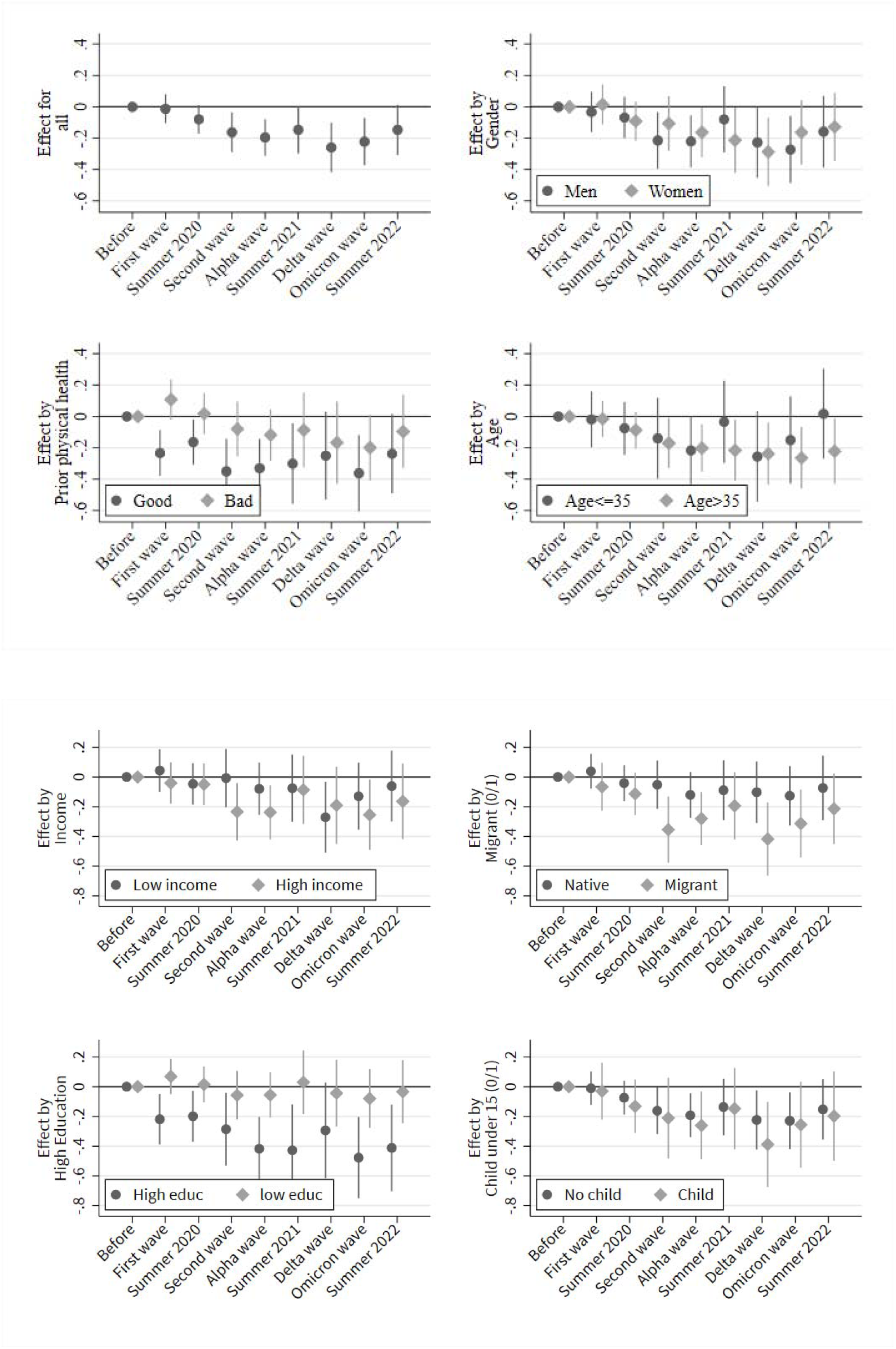
Effects on psychological well-being.

**Figure A3.**
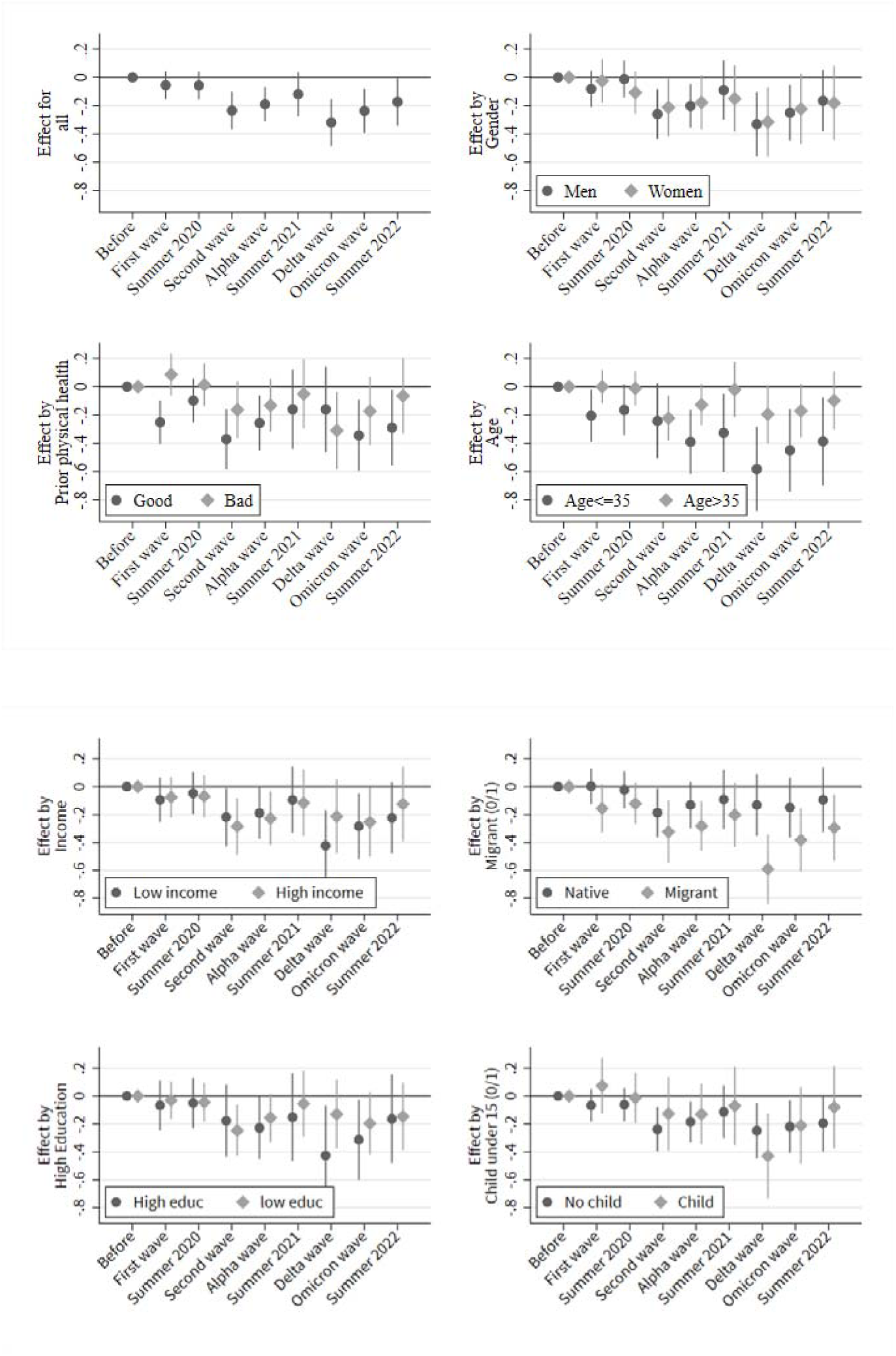
Effects on feeling calm.

**Figure A4.**
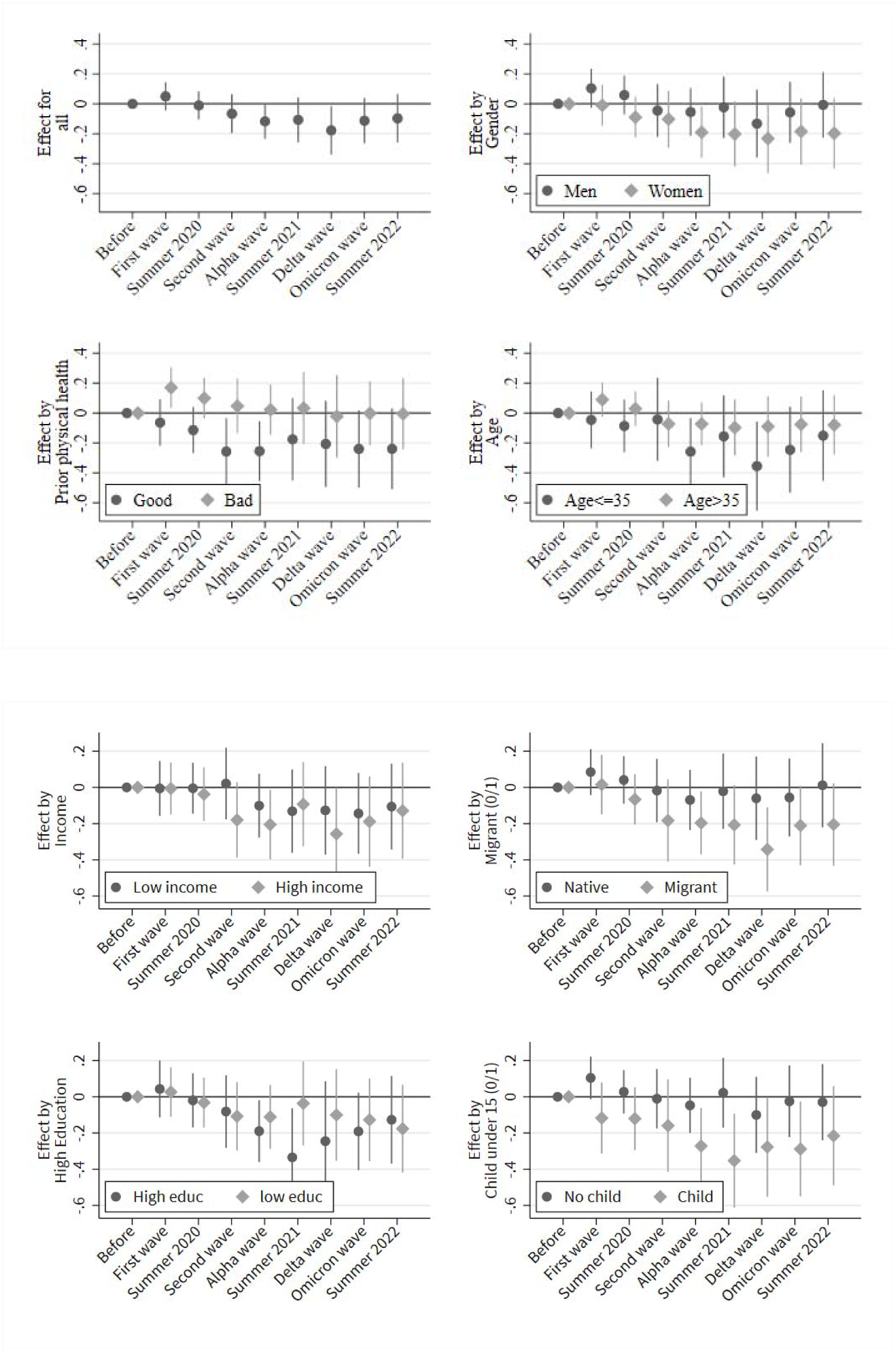
Effects on vitality.

**Figure A5.**
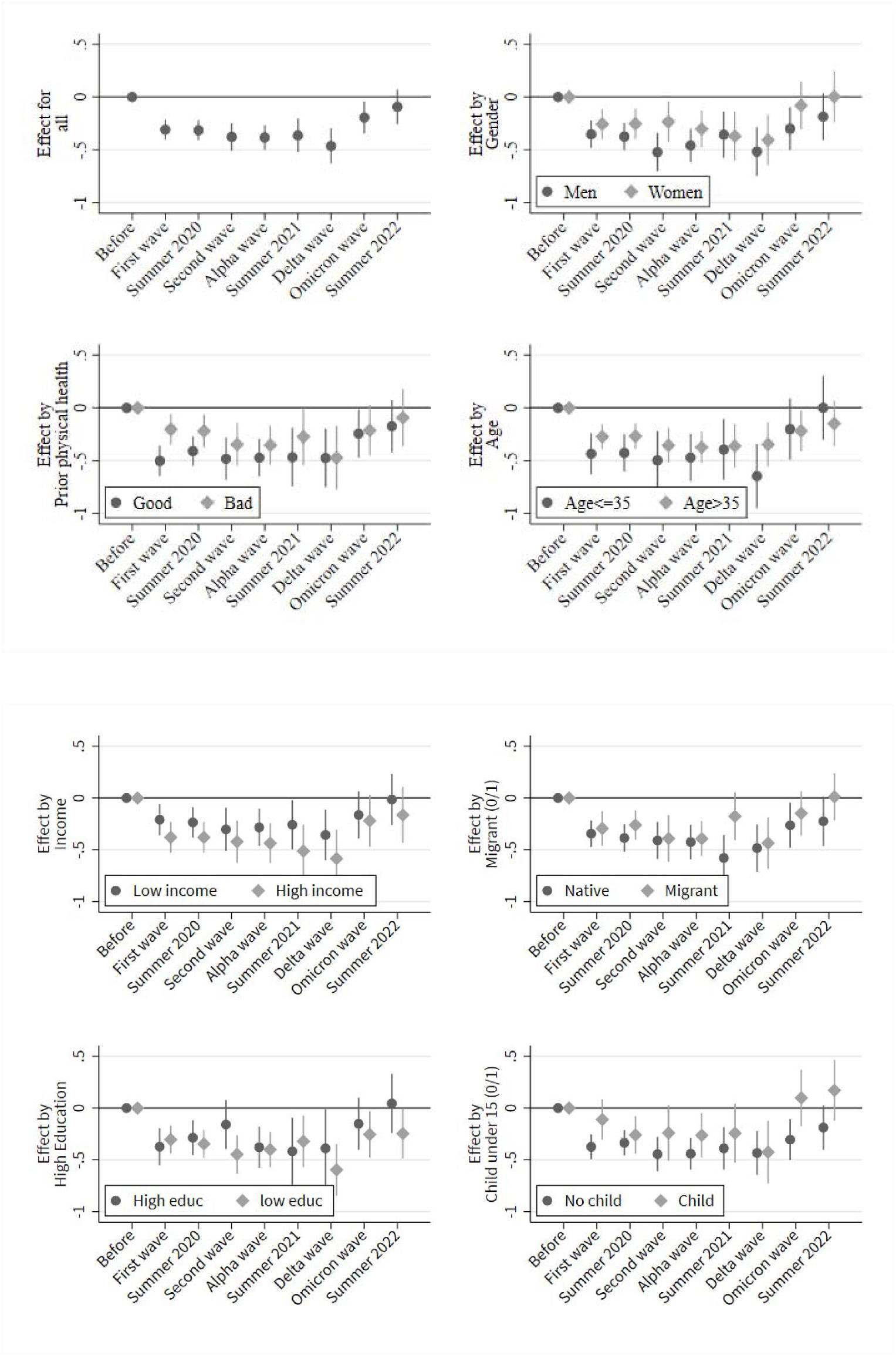
Effects on emotional stability.

**Table A1:** Results corresponding to Figure 3

|  | (1) | (2) | (3) | (4) | (5) | (6) | (7) |
| --- | --- | --- | --- | --- | --- | --- | --- |
|  | Main | Men | Women | Good health | Bad health | Age <35 | Age ≥35 |
| **Pandemic phase** |  |  |  |  |  |  |  |
| First wave<br>(baseline: before) | -0.081<sup>*</sup><br>(0.032) | -0.089<sup>*</sup><br>(0.043) | -0.070<br>(0.049) | -0.264<sup>***</sup><br>(0.050) | 0.048<br>(0.048) | -0.178<sup>**</sup><br>(0.061) | -0.048<br>(0.040) |
| Summer 2020 | -0.114<sup>***</sup><br>(0.032) | -0.097<sup>*</sup><br>(0.043) | -0.134<sup>**</sup><br>(0.049) | -0.197<sup>***</sup><br>(0.049) | -0.016<br>(0.049) | -0.186<sup>**</sup><br>(0.057) | -0.083<sup>*</sup><br>(0.041) |
| Second wave | -0.212<sup>***</sup><br>(0.044) | -0.265<sup>***</sup><br>(0.060) | -0.163<sup>*</sup><br>(0.066) | -0.369<sup>***</sup><br>(0.071) | -0.131<sup>*</sup><br>(0.066) | -0.242<sup>**</sup><br>(0.088) | -0.205<sup>***</sup><br>(0.055) |
| Alpha wave | -0.221<sup>***</sup><br>(0.040) | -0.233<sup>***</sup><br>(0.053) | -0.209<sup>***</sup><br>(0.060) | -0.328<sup>***</sup><br>(0.063) | -0.139<sup>*</sup><br>(0.061) | -0.334<sup>***</sup><br>(0.071) | -0.194<sup>***</sup><br>(0.051) |
| Summer 2021 | -0.179<sup>***</sup><br>(0.052) | -0.132<br>(0.071) | -0.229<sup>**</sup><br>(0.076) | -0.278<sup>**</sup><br>(0.089) | -0.077<br>(0.085) | -0.223<sup>*</sup><br>(0.088) | -0.168<sup>*</sup><br>(0.067) |
| Delta wave | -0.299<sup>***</sup><br>(0.056) | -0.291<sup>***</sup><br>(0.075) | -0.308<sup>***</sup><br>(0.084) | -0.279<sup>**</sup><br>(0.097) | -0.224<sup>*</sup><br>(0.100) | -0.452<sup>***</sup><br>(0.098) | -0.211<sup>**</sup><br>(0.070) |
| Omicron wave | -0.191<sup>***</sup><br>(0.051) | -0.221<sup>**</sup><br>(0.068) | -0.162<sup>*</sup><br>(0.078) | -0.299<sup>***</sup><br>(0.082) | -0.138<br>(0.078) | -0.264<sup>**</sup><br>(0.091) | -0.182<sup>**</sup><br>(0.065) |
| Summer 2022 | -0.125<sup>*</sup><br>(0.055) | -0.130<br>(0.074) | -0.119<br>(0.083) | -0.234<sup>**</sup><br>(0.087) | -0.055<br>(0.086) | -0.128<br>(0.095) | -0.134<br>(0.070) |
| **Interview month** |  |  |  |  |  |  |  |
| March (baseline:<br>February) | -0.020<br>(0.017) | -0.013<br>(0.024) | -0.027<br>(0.025) | -0.023<br>(0.027) | -0.033<br>(0.024) | 0.067<br>(0.044) | -0.040<sup>*</sup><br>(0.019) |
| April | -0.008<br>(0.020) | -0.014<br>(0.027) | -0.004<br>(0.028) | 0.005<br>(0.031) | -0.026<br>(0.028) | 0.090<br>(0.047) | -0.035<br>(0.022) |
| May | -0.023<br>(0.021) | -0.042<br>(0.029) | -0.005<br>(0.031) | -0.029<br>(0.033) | -0.041<br>(0.031) | 0.060<br>(0.049) | -0.049<sup>*</sup><br>(0.024) |
| June | -0.026<br>(0.026) | -0.071*<br>(0.036) | 0.019<br>(0.038) | -0.085*<br>(0.040) | -0.042<br>(0.040) | 0.073<br>(0.056) | -0.059<br>(0.032) |
| July | 0.007<br>(0.029) | -0.043<br>(0.039) | 0.059<br>(0.043) | -0.009<br>(0.043) | -0.047<br>(0.044) | 0.098<br>(0.059) | -0.034<br>(0.036) |
| August | -0.014<br>(0.031) | -0.051<br>(0.043) | 0.025<br>(0.046) | -0.060<br>(0.049) | -0.054<br>(0.049) | 0.071<br>(0.064) | -0.048<br>(0.039) |
| September | -0.003<br>(0.036) | -0.018<br>(0.050) | 0.013<br>(0.054) | -0.040<br>(0.058) | -0.016<br>(0.061) | 0.110<br>(0.071) | -0.053<br>(0.045) |
| **Interview mode** |  |  |  |  |  |  |  |
| CAPI, face-to-face<br>(baseline: CATI) | -0.003<br>(0.019) | -0.017<br>(0.026) | 0.009<br>(0.028) | 0.044<br>(0.028) | -0.032<br>(0.027) | -0.028<br>(0.038) | 0.009<br>(0.023) |
| CAPI, phone | 0.023<br>(0.015) | 0.008<br>(0.021) | 0.040<br>(0.022) | 0.022<br>(0.024) | 0.039<br>(0.024) | 0.022<br>(0.030) | 0.036<br>(0.019) |
| Age (years) | 0.015<br>(0.012) | 0.022<br>(0.016) | 0.009<br>(0.019) | 0.007<br>(0.020) | 0.021<br>(0.019) | 0.023<br>(0.021) | 0.016<br>(0.016) |
| Constant | 2.929***<br>(0.542) | 2.755***<br>(0.715) | 3.126***<br>(0.838) | 3.757***<br>(0.851) | 2.294*<br>(0.937) | 3.012***<br>(0.530) | 2.773***<br>(0.841) |
| Observations | 40020 | 20058 | 19956 | 13158 | 17565 | 12534 | 27486 |
| Individuals | 18704 | 9608 | 9100 | 5793 | 7171 | 7305 | 12055 |

Table A 1: Results corresponding to Figure 3 (cont)
|  | (1) | (2) | (3) | (4) | (5) | (6) | (7) | (8) |
| --- | --- | --- | --- | --- | --- | --- | --- | --- |
|  | Low<br>income | High<br>income | Native | Migrant | High<br>education | Low<br>education | No<br>child | Children |
| **Pandemic phase** |  |  |  |  |  |  |  |  |
| First wave (baseline:<br>before) | -0.062<br>(0.049) | -0.121*<br>(0.050) | -0.053<br>(0.044) | -0.126*<br>(0.056) | -0.055<br>(0.046) | -0.153**<br>(0.053) | -0.085*<br>(0.040) | -0.046<br>(0.067) |
| Summer 2020 | -0.081<br>(0.047) | -0.131*<br>(0.052) | -0.100*<br>(0.046) | -0.139**<br>(0.047) | -0.097*<br>(0.047) | -0.140**<br>(0.052) | -0.110**<br>(0.041) | -0.130*<br>(0.061) |
| Second wave | -0.132*<br>(0.065) | -0.275***<br>(0.071) | -0.163**<br>(0.061) | -0.322***<br>(0.076) | -0.212***<br>(0.063) | -0.177*<br>(0.074) | -0.216***<br>(0.056) | -0.183*<br>(0.090) |
| Alpha wave | -0.164**<br>(0.057) | -0.272***<br>(0.066) | -0.184**<br>(0.058) | -0.290***<br>(0.056) | -0.175**<br>(0.059) | -0.304***<br>(0.061) | -0.217***<br>(0.052) | -0.229**<br>(0.074) |
| Summer 2021 | -0.134<br>(0.074) | -0.198*<br>(0.082) | -0.192**<br>(0.074) | -0.190**<br>(0.073) | -0.086<br>(0.079) | -0.325***<br>(0.096) | -0.150*<br>(0.066) | -0.196*<br>(0.092) |
| Delta wave | -0.287***<br>(0.081) | -0.307**<br>(0.096) | -0.192*<br>(0.079) | -0.440***<br>(0.080) | -0.204*<br>(0.085) | -0.339**<br>(0.122) | -0.249***<br>(0.070) | -0.371***<br>(0.103) |
| Omicron wave | -0.181*<br>(0.072) | -0.224**<br>(0.086) | -0.146<br>(0.075) | -0.266***<br>(0.070) | -0.158*<br>(0.076) | -0.285***<br>(0.076) | -0.196**<br>(0.067) | -0.161<br>(0.092) |
| Summer 2022 | -0.097<br>(0.077) | -0.140<br>(0.092) | -0.092<br>(0.082) | -0.173*<br>(0.074) | -0.143<br>(0.081) | -0.157<br>(0.087) | -0.140*<br>(0.071) | -0.073<br>(0.099) |
| **Interview month** |  |  |  |  |  |  |  |  |
| March (baseline:<br>February) | 0.000<br>(0.028) | -0.047*<br>(0.024) | -0.018<br>(0.019) | -0.030<br>(0.042) | -0.044*<br>(0.021) | 0.059<br>(0.038) | -0.028<br>(0.019) | 0.012<br>(0.047) |
| April | 0.013<br>(0.033) | -0.019<br>(0.027) | -0.000<br>(0.022) | -0.046<br>(0.046) | -0.021<br>(0.024) | 0.059<br>(0.042) | -0.023<br>(0.022) | 0.038<br>(0.051) |
| May | -0.038<br>(0.034) | -0.015<br>(0.030) | -0.011<br>(0.025) | -0.065<br>(0.046) | -0.026<br>(0.026) | -0.006<br>(0.046) | -0.032<br>(0.024) | 0.016<br>(0.051) |
| June | -0.004<br>(0.042) | -0.080*<br>(0.038) | -0.014<br>(0.032) | -0.066<br>(0.052) | -0.029<br>(0.034) | -0.029<br>(0.054) | -0.043<br>(0.031) | 0.045<br>(0.059) |
| July | -0.050<br>(0.046) | 0.036<br>(0.042) | 0.041<br>(0.036) | -0.057<br>(0.055) | -0.022<br>(0.038) | 0.082<br>(0.058) | 0.002<br>(0.035) | 0.052<br>(0.064) |
| August | 0.023<br>(0.050) | -0.071<br>(0.046) | -0.005<br>(0.040) | -0.063<br>(0.059) | -0.028<br>(0.042) | 0.037<br>(0.066) | -0.051<br>(0.037) | 0.074<br>(0.069) |
| September | -0.000<br>(0.055) | -0.067<br>(0.058) | 0.013<br>(0.048) | -0.059<br>(0.064) | 0.005<br>(0.051) | -0.029<br>(0.086) | -0.020<br>(0.044) | 0.055<br>(0.077) |
| **Interview mode** |  |  |  |  |  |  |  |  |
| CAPI, face-to-face<br>(baseline: CATI) | -0.005<br>(0.030) | 0.007<br>(0.028) | 0.001<br>(0.024) | -0.009<br>(0.033) | -0.010<br>(0.025) | 0.014<br>(0.039) | -0.016<br>(0.023) | 0.011<br>(0.040) |
| CAPI, phone | -0.000<br>(0.024) | 0.042<br>(0.023) | 0.024<br>(0.020) | 0.044<br>(0.027) | 0.028<br>(0.020) | 0.008<br>(0.033) | 0.018<br>(0.019) | 0.056<br>(0.030) |
| Age (years) | 0.020<br>(0.017) | 0.017<br>(0.021) | 0.006<br>(0.018) | 0.023<br>(0.016) | 0.007<br>(0.018) | 0.036*<br>(0.017) | 0.017<br>(0.016) | 0.000<br>(0.021) |
| Constant | 2.582***<br>(0.721) | 2.993**<br>(0.956) | 3.281***<br>(0.870) | 2.815***<br>(0.623) | 3.253***<br>(0.891) | 1.978*<br>(0.825) | 2.774***<br>(0.766) | 3.629***<br>(0.748) |
| Observations | 20305 | 19237 | 25216 | 14010 | 22097 | 6026 | 29633 | 10387 |
| Individuals | 11935 | 9207 | 10730 | 7609 | 8433 | 2122 | 13809 | 5750 |

**Table A2:** Results corresponding to Figure 4.

|  | (1) | (2) | (3) | (4) |
| --- | --- | --- | --- | --- |
|  | Psychological Well-being | Feeling Calm | Vitality | Role emotional |
| **Pandemic phase** |  |  |  |  |
| First wave (baseline:<br>before) | -0.013<br>(0.047) | -0.055<br>(0.050) | 0.050<br>(0.048) | -0.309***<br>(0.049) |
| Summer 2020 | -0.080<br>(0.047) | -0.057<br>(0.050) | -0.010<br>(0.048) | -0.315***<br>(0.049) |
| Second wave | -0.162*<br>(0.065) | -0.235***<br>(0.068) | -0.066<br>(0.066) | -0.379***<br>(0.067) |
| Alpha wave | -0.196**<br>(0.060) | -0.190**<br>(0.062) | -0.117*<br>(0.060) | -0.384***<br>(0.060) |
| Summer 2021 | -0.147<br>(0.076) | -0.119<br>(0.080) | -0.107<br>(0.076) | -0.365***<br>(0.081) |
| Delta wave | -0.259**<br>(0.081) | -0.319***<br>(0.085) | -0.177*<br>(0.083) | -0.465***<br>(0.085) |
| Omicron wave | -0.222**<br>(0.077) | -0.237**<br>(0.080) | -0.113<br>(0.077) | -0.195*<br>(0.077) |
| Summer 2022 | -0.147<br>(0.082) | -0.173*<br>(0.086) | -0.097<br>(0.082) | -0.094<br>(0.083) |
| **Interview month** |  |  |  |  |
| March (baseline:<br>February) | -0.010<br>(0.023) | -0.043<br>(0.026) | 0.003<br>(0.025) | -0.027<br>(0.025) |
| April | -0.026<br>(0.026) | -0.037<br>(0.029) | 0.036<br>(0.028) | 0.001<br>(0.029) |
| May | -0.016<br>(0.029) | -0.053<br>(0.031) | 0.030<br>(0.031) | -0.047<br>(0.031) |
| June | -0.025<br>(0.036) | -0.064<br>(0.038) | 0.058<br>(0.038) | -0.068<br>(0.039) |
| July | 0.032<br>(0.040) | -0.019<br>(0.043) | 0.079<br>(0.042) | -0.045<br>(0.043) |
| August | 0.007<br>(0.043) | -0.064<br>(0.046) | 0.059<br>(0.047) | -0.042<br>(0.047) |
| September | -0.008<br>(0.051) | -0.059<br>(0.056) | 0.119*<br>(0.054) | -0.051<br>(0.057) |
| **Interview mode** |  |  |  |  |
| CAPI, face-to-face<br>(baseline: CATI) | 0.064*<br>(0.026) | -0.086**<br>(0.028) | -0.063*<br>(0.028) | 0.063*<br>(0.028) |
| CAPI, phone | 0.062**<br>(0.021) | -0.012<br>(0.023) | -0.032<br>(0.023) | 0.069**<br>(0.024) |
| Age (years) | 0.017<br>(0.019) | 0.020<br>(0.019) | 0.006<br>(0.019) | 0.017<br>(0.019) |
| Constant | 2.923***<br>(0.825) | 2.641**<br>(0.854) | 2.805***<br>(0.822) | 3.412***<br>(0.824) |
| Observations | 40285 | 40268 | 40223 | 40146 |
| Individuals | 18833 | 18825 | 18803 | 18755 |

Standard errors in parentheses
\* $p < 0.05$ , \*\* $p < 0.01$ , \*\*\* $p < 0.001$

