## Supplementary material for "Mental health in Germany before, during and after the COVID-19 pandemic": Replication Syntax: PLOS_One_Covid_mental_health.do

```
/*
Replication Syntax for the Paper:
"Mental health in Germany before, during and after the COVID-19 pandemic"

Authors:
Alexander Patzina, Matthias Collischon, Rasmus Hoffmann, Maksym Obrizan

published in PLOS One
*/

*********Data Prep**************

cd "[PASS data Directory]"


global results "[enter your results folder]"

adopath + "[ado path]"


***Load the data
use "PENDDAT.dta", clear


***merge household data
merge m:1 hnr welle using "HHENDDAT.dta", gen(_hmerge)
keep if _hmerge==3


**Generate interview date: Year/Month
gen pintmonyear=ym(pintjahr, pintmon)
format pintmonyear %tm
label var pintmonyear "Year/month"


**Drop observations prior to 2018
keep if welle>=12
count


***generate exact interview date
gen intdat=mdy(pintmon,pinttag, pintjahr)
format intdat %td

*week
cap gen week_num=.
foreach y of numlist 2014(1)2022 {
    replace  week_num=wofd(intdat)-wofd(td(31dec`y')) if pintjahr==`y'+1
}


***Outcome: mental health score
tab1 PG1215*
foreach i of varlist PG1215a PG1215b PG1215c PG1215g {
    recode `i' (-99/-1=.)(6/.=.), gen(`i'_rec)
	
}

tab1 PG1215*_rec
**Cronbach's alpha: 0.77
alpha  PG1215*_rec

***reverse
foreach i of varlist  PG1215b_rec PG1215c_rec   { //PG1215g_rec
recode `i' (1=5)(2=4)(4=2)(5=1)
}


*Factor Analysis
factor PG1215*_rec 
*Eigenvalue=1.76

**Generate mental health Index
egen index_mental_health=rowmean(PG1215*_rec )
label var index_mental_health "Mental health"
sum index_mental_health, d 

**replace values if missing
foreach y of varlist PG1215*_rec {
    replace index_mental_health=. if `y'==.
    
}

***Subgroups
tab1 PG1200 
*Health in 2018
gen good_phealth18h = inlist(PG1200,1,2) if welle==12
bysort pnr: egen good_phealth18 = max(good_phealth18h)
tab good_phealth18
label var good_phealth18 "Good health"


****Migration
gen migrant=.
replace migrant=1 if migration>1
replace migrant=0 if migration==1
label var migrant "Migrant (0/1)"


***Education
numlabel beruf2, add
tab beruf2
	
gen 	educ_voc = 0 if inlist(beruf2,2,3)
replace educ_voc = 1 if inlist(beruf2,4,5,6,7,8,12,13)
replace educ_voc = 2 if inlist(beruf2,9,10,11)
	
gen educ_voc13h = educ_voc if welle==13
by pnr, sort: egen educ_voc13=max(educ_voc13h)
	
tab educ_voc13

gen lowqual13 = educ_voc13==0 if inlist(educ_voc13,0,1,2) 
tab lowqual13

gen highqual13 = educ_voc13==2 if inlist(educ_voc13,0,1,2) 
tab highqual13
	
label var highqual "High Education"

	
***Child u15
gen kid=(kindu15==1)
label var kid "Child under 15 (0/1)"
	
****Other outcomes
****Health satisfaction
tab PA0100
recode PA0100 (-99/-1=.)(11/9999=.), gen(healthsat)
label var healthsat "Health satisfaction"

*************************************************
****Subjective health
tab PG1200
recode PG1200 (-99/-1=.)(11/9999=.)(1=5)(2=4)(4=2)(5=1), gen(ovr_sub_health) //reverse coded, code so that higher=better
label var ovr_sub_health "Subjective health"
*Binary
recode PG1200 (1 2=1)(3 4 5=0) (-99/0=.), gen(ovr_good_health)
label var ovr_good_health "Good health (0/1)"

*****Psych health
tab PG1100
recode PG1100 (-99/-1=.)(11/9999=.)(1=5)(2=4)(4=2)(5=1), gen(sub_psy_health) //reverse coded, code so that higher=better
label var sub_psy_health "Mental health"
*Binary
recode PG1100 (1 2=1)(3 4 5=0) (-99/0=.), gen(good_psy_health)
label var good_psy_health "Few mental health problems (0/1)"

***Subscales as observed
label var PG1215a_rec "Psychological Well-being" //"Feeling depressed (PG1215a)"
label var PG1215b_rec "Feeling calm"
label var PG1215c_rec "Vitality"
label var PG1215g_rec "Emotional stability"


*Sample restrictions
keep if healthsat!=. & sub_psy_health!=.

global outcomes "healthsat ovr_sub_health ovr_good_health sub_psy_health good_psy_health index_mental_health PG1215a_rec  PG1215b_rec PG1215c_rec PG1215g_rec"


****Results by group
gen female=.
replace female=1 if zpsex==2 
replace female=0 if zpsex==1
label var female "Gender"
label def female 0 "Men" 1 "Women"
label val female female

**Age
cap drop young
gen young=.
replace young=0 if palter <=35
replace young=1 if palter >35
label var young "Age"
label def young 0 "Age <=35" 1 "Age>35"
label val young young


***Good health in 2018
label var good_phealth18 "Physical health 2018"
label def good_phealth18 0 "Bad" 1 "Good"
label val good_phealth18 good_phealth18


cap drop high_inc
gen high_inc=.
sum oecdincn if oecdincn>0 & pintjahr==2020, d
replace high_inc=0 if oecdincn<=r(p50) & oecdincn>0
replace high_inc=1 if oecdincn>r(p50) & oecdincn!=.
gen eq_inc=oecdincn if oecdincn>0 
label var eq_inc "Needs-adjusted household income"
lab var high_inc "Income"


***Generate variables for event study

****Event study - which wave?
cap drop wave
gen wave=.
replace wave=0 if intdat<=mdy(31,1,2020)
replace wave=1 if intdat>=mdy(2,1,2020) & intdat<=mdy(5,17,2020)
replace wave=2 if intdat>=mdy(5,18,2020) & intdat<=mdy(9,27,2020)
replace wave=3 if intdat>=mdy(9,28,2020) & intdat<=mdy(2,28,2021)
replace wave=4 if intdat>=mdy(3,1,2021) & intdat<=mdy(6,13,2021)
replace wave=5 if intdat>=mdy(6,13,2021) & intdat<=mdy(8,1,2021)
replace wave=6 if intdat>=mdy(8,2,2021) & intdat<=mdy(12,26,2021)
replace wave=7 if intdat>=mdy(12,27,2021) & intdat<=mdy(5,24,2022)
replace wave=8 if intdat>=mdy(5,25,2022)
label var wave "Pandemic phase"
label def wave 0 "Before" 1 "First wave" 2 "Summer 2020" 3 "Second wave" 4 "Alpha wave" 5 "Summer 2021" 6 "Delta wave" 7 "Omicron wave" 8 "Summer 2022", replace
label val wave wave


***Generate output for Table 1
tab wave, gen(wave_)


cap gen ost=.
replace ost=1 if blneualt==2
replace ost=0 if blneualt==1
label var ost "East Germany (0/1)"
cap tab educ_voc, gen(educ_)
label var educ_1 "No vocational or university degree"
label var educ_2 "Vocational  degree"
label var educ_3 "University degree"
label var good_phealth18 "Prior physical health"
label var palter "Age (years)"

gen cati=(pintmod==0)
label var cati "CATI"
gen capi=(pintmod==1)
label var capi "CAPI, face to face"
gen capi2=(pintmod==2)
label var capi2 "CAPI, by phone"

global desc "healthsat index_mental_health PG1215*_rec sub_psy_health  female palter good_phealth18  pintmon cati capi capi2  wave_* migrant high_inc kid highqual13"

estpost summarize $desc if sub_psy_health!=. & healthsat!=.
eststo othersum


***Before/after Pandemic

*****Table 2 output
****Samples
estpost summarize $desc if index_mental_health!=. &  sub_psy_health!=. & healthsat!=.
eststo indexsum

estpost summarize $desc if pintjahr<=2019 & index_mental_health!=. &  sub_psy_health!=. & healthsat!=.
eststo sum2019

estpost summarize $desc if pintjahr>=2020 & index_mental_health!=. &  sub_psy_health!=. & healthsat!=.
eststo sum2020

esttab indexsum sum2019 sum2020 using "$results/descriptives_time.rtf" ///
		, cells("mean(fmt(%9.2f) pattern(1 1 1 1)) sd(fmt(%9.2f) pattern(1 1 1 1))  ") ///
label    replace nogaps      compress


****Figure 3-1  + Table A1 and Figures A2 to A5, upper parts
est clear
global controls " i.pintmon i.pintmod c.palter"
foreach y of varlist healthsat   sub_psy_health good_psy_health  PG1215*_rec index_mental_health {
local label: variable label `y'
xtreg `y' i.wave $controls, cl(pnr) fe
est sto main
estadd scalar obs=e(N)
estadd scalar people=e(N_clust)
margins,  dydx(wave) post
coefplot .,   scheme(sj) graphr(color(white)) xlab(, angle(45))   ytitle("Effect for" "all") base vert yline(0)  keep(*av*)
graph save "$results/`y'_main.gph", replace


****By gender
local label_sub: variable label female
xtreg `y' i.wave  $controls if female==0, cl(pnr) fe
est sto male
estadd scalar obs=e(N)
estadd scalar people=e(N_clust)
xtreg `y' i.wave  $controls if female==1, cl(pnr) fe
est sto female
estadd scalar obs=e(N)
estadd scalar people=e(N_clust)

coefplot (male, label("Men")) (female, label("Women")),  scheme(sj) graphr(color(white)) xlab(, angle(45))  ytitle("Effect by" "`label_sub'") base vert yline(0)  keep(*av*) legend(ring(0) position(8))
graph save "$results/`y'_gender.gph", replace

***By migration
local label_sub: variable label migrant
xtreg `y' i.wave  $controls if migrant==0, cl(pnr) fe
est sto native
estadd scalar obs=e(N)
estadd scalar people=e(N_clust)
xtreg `y' i.wave  $controls if migrant==1, cl(pnr) fe
est sto migrant
estadd scalar obs=e(N)
estadd scalar people=e(N_clust)

coefplot (native, label("Native")) (migrant, label("Migrant")),  scheme(sj) graphr(color(white)) xlab(, angle(45))  ytitle("Effect by" "`label_sub'") base vert yline(0)  keep(*av*) legend(ring(0) position(8))
graph save "$results/`y'_migrant.gph", replace
graph export "$results/`y'_migrant.png", replace


*By Health in 2018
local label_sub: variable label good_phealth18
xtreg `y' i.wave  $controls if good_phealth18==0, cl(pnr) fe
est sto bhealth
estadd scalar obs=e(N)
estadd scalar people=e(N_clust)
xtreg `y' i.wave  $controls if good_phealth18==1, cl(pnr) fe
est sto ghealth
estadd scalar obs=e(N)
estadd scalar people=e(N_clust)

coefplot (ghealth, label("Good")) (bhealth, label("Bad")),    scheme(sj) graphr(color(white)) xlab(, angle(45))  ytitle("Effect by" "`label_sub'") base vert yline(0) keep(*av*) legend(ring(0) position(8))
graph save "$results/`y'_health2018.gph", replace


*By age 
local label_sub: variable label young
xtreg `y' i.wave  $controls if young==0, cl(pnr) fe
est sto young
estadd scalar obs=e(N)
estadd scalar people=e(N_clust)
xtreg `y' i.wave  $controls if young==1, cl(pnr) fe
est sto old
estadd scalar obs=e(N)
estadd scalar people=e(N_clust)

coefplot (young, label("Age<=35")) (old, label("Age>35")),   scheme(sj) graphr(color(white)) xlab(, angle(45)) ytitle("Effect by" "`label_sub'") base vert yline(0)  keep(*av*) legend(ring(0) position(8))
graph save "$results/`y'_young.gph", replace

esttab main male female ghealth bhealth young old ,  b(%9.3f) se(%9.3f) stats(obs people, fmt(%6.0f) labels("Observations" "Individuals")) mti label
esttab main male female ghealth bhealth young old using "$results/`y'_reg.rtf", replace b(%9.3f) se(%9.3f) stats(obs people, fmt(%6.0f) labels("Observations" "Individuals")) mti label

***Combine
graph combine  "$results/`y'_main.gph" "$results/`y'_gender.gph" "$results/`y'_health2018.gph" "$results/`y'_young.gph", ycommon xcommon c(2) graphr(color(white)) // note("Individual FE-Regressions. Conditional on: Interview month, interview mode, age (linear).") //title("`label'")

graph export "$results/`y'_wave.png", replace
graph export "$results/`y'_wave.pdf", replace
graph export "$results/`y'_wave.wmf", replace
}


***Loop creates Figure 3_2 + Table A1 and Figures A2 to A5, lower parts
est clear
global controls " i.pintmon i.pintmod c.palter"
foreach y of varlist index_mental_health {
local label: variable label `y'


***By Income
local label_sub: variable label high_inc
xtreg `y' i.wave  $controls if high_inc==0, cl(pnr) fe
est sto linc
estadd scalar obs=e(N)
estadd scalar people=e(N_clust)
xtreg `y' i.wave  $controls if high_inc==1, cl(pnr) fe
est sto hinc
estadd scalar obs=e(N)
estadd scalar people=e(N_clust)

coefplot (linc, label("Low income")) (hinc, label("High income")),  scheme(sj) graphr(color(white)) xlab(, angle(45))  ytitle("Effect by" "`label_sub'") base vert yline(0)  keep(*av*) legend(ring(0) position(8))
graph save "$results/`y'_hinc2020.gph", replace
graph export "$results/`y'_hinc2020.png", replace


***By migration
local label_sub: variable label migrant
xtreg `y' i.wave  $controls if migrant==0, cl(pnr) fe
est sto native
estadd scalar obs=e(N)
estadd scalar people=e(N_clust)
xtreg `y' i.wave  $controls if migrant==1, cl(pnr) fe
est sto migrant
estadd scalar obs=e(N)
estadd scalar people=e(N_clust)

coefplot (native, label("Native")) (migrant, label("Migrant")),  scheme(sj) graphr(color(white)) xlab(, angle(45))  ytitle("Effect by" "`label_sub'") base vert yline(0)  keep(*av*) legend(ring(0) position(8))
graph save "$results/`y'_migrant.gph", replace
graph export "$results/`y'_migrant.png", replace


*By Education
local label_sub: variable label highqual13
xtreg `y' i.wave  $controls if highqual13==0, cl(pnr) fe
est sto beduc
estadd scalar obs=e(N)
estadd scalar people=e(N_clust)
xtreg `y' i.wave  $controls if highqual13==1, cl(pnr) fe
est sto geduc
estadd scalar obs=e(N)
estadd scalar people=e(N_clust)

coefplot (geduc, label("High educ")) (beduc, label("low educ")),    scheme(sj) graphr(color(white)) xlab(, angle(45))  ytitle("Effect by" "`label_sub'") base vert yline(0) keep(*av*) legend(ring(0) position(8))
graph save "$results/`y'_educ2019.gph", replace


*By children 
local label_sub: variable label kid
xtreg `y' i.wave  $controls if kid==0, cl(pnr) fe
est sto nokid
estadd scalar obs=e(N)
estadd scalar people=e(N_clust)
xtreg `y' i.wave  $controls if kid==1, cl(pnr) fe
est sto kid
estadd scalar obs=e(N)
estadd scalar people=e(N_clust)

coefplot (nokid, label("No child")) (kid, label("Child")),   scheme(sj) graphr(color(white)) xlab(, angle(45)) ytitle("Effect by" "`label_sub'") base vert yline(0)  keep(*av*) legend(ring(0) position(8))
graph save "$results/`y'_kid.gph", replace

esttab linc hinc native migrant beduc geduc nokid kid ,  b(%9.3f) se(%9.3f) stats(obs people, fmt(%6.0f) labels("Observations" "Individuals")) mti label
esttab linc hinc  native migrant beduc geduc nokid kid using "$results/`y'_reg2.rtf", replace b(%9.3f) se(%9.3f) stats(obs people, fmt(%6.0f) labels("Observations" "Individuals")) mti label

***Combine
graph combine  "$results/`y'_hinc2020.gph" "$results/`y'_migrant.gph" "$results/`y'_educ2019.gph" "$results/`y'_kid.gph", ycommon xcommon c(2) graphr(color(white)) // note("Individual FE-Regressions. Conditional on: Interview month, interview mode, age (linear).") //title("`label'")

graph export "$results/`y'_wave2.png", replace
graph export "$results/`y'_wave2.pdf", replace
graph export "$results/`y'_wave2.wmf", replace
}


***create Figure 4 and Table A2
est clear
global controls " i.pintmon i.pintmod c.palter"
foreach y of varlist PG1215*_rec {
local label: variable label `y'
xtreg `y' i.wave $controls, cl(pnr) fe
est sto `y'_main 
estadd scalar obs=e(N)
estadd scalar people=e(N_clust)
margins,  dydx(wave) post
coefplot .,   scheme(sj) graphr(color(white)) xlab(, angle(45))   ytitle("Effect on" "`label'") base vert yline(0)  keep(*av*) xtitle("Pandemic phase")
graph save "$results/`y'_main_lab.gph", replace
}

esttab *_main ,  b(%9.3f) se(%9.3f) stats(obs people, fmt(%6.0f) labels("Observations" "Individuals")) mti label
esttab *_main using "$results/subscores_reg.rtf", replace b(%9.3f) se(%9.3f) stats(obs people, fmt(%6.0f) labels("Observations" "Individuals")) mti label


graph combine  "$results/PG1215a_rec_main_lab.gph" "$results/PG1215b_rec_main_lab.gph" "$results/PG1215c_rec_main_lab.gph" "$results/PG1215g_rec_main_lab.gph", ycommon xcommon c(2) graphr(color(white)) // note("Individual FE-Regressions. Conditional on: Interview month, interview mode, age (linear).") 

graph export "$results/sf12_wave_comb.png", replace
graph export "$results/`y'sf12_wave_comb.pdf", replace
graph export "$results/`y'sf12_wave_comb.wmf", replace


***Effects on other outcomes - used for Figure A1
est clear
global controls " i.pintmon i.pintmod c.palter"
foreach y of varlist healthsat  sub_psy_health ovr_sub_health{
local label: variable label `y'
xtreg `y' i.wave $controls, cl(pnr) fe
margins,  dydx(wave) post
coefplot .,   scheme(sj) graphr(color(white)) xlab(, angle(45))   ytitle("Effect for" "all") base vert yline(0)  keep(*av*)  title("`label'")
graph save "$results/`y'_mainx.gph", replace


}


***Mean plots over time
***Loop generates Figure 1
foreach y of varlist healthsat  index_mental_health  sub_psy_health good_psy_health PG1215a_rec PG1215b_rec PG1215c_rec PG1215g_rec  {
local label: variable label `y' 
global healthsat_lab "5(1)8"
global index_mental_health_lab "3(0.25)4"
global sub_psy_health_lab "3(0.25)4"
preserve

bysort wave: egen m_`y'=mean(`y')
bysort wave: keep if _n==1
disp "Graph"
twoway (connected m_`y' wave , mcol(black)),  scheme(sj) graphr(color(white)) xlab(0(1)8, angle(45) valuelabel)  ytitle("`label'") ///
 legend(off) ylab($`y'_lab)
graph save "$results/`y'_desc_waves.gph", replace
graph export "$results/`y'_desc_waves.png", replace
restore
}


***Figure 2
graph combine  "$results/PG1215a_rec_desc_waves.gph" "$results/PG1215b_rec_desc_waves.gph" "$results/PG1215c_rec_desc_waves.gph" "$results/PG1215g_rec_desc_waves.gph", ycommon xcommon c(2) graphr(color(white))   

graph export "$results/sf12_wave_desc.png", replace
graph export "$results/sf12_wave_desc.pdf", replace
graph export "$results/sf12_wave_desc.wmf", replace


***Figure A1
graph combine  "$results/healthsat_mainx.gph" "$results/sub_psy_health_mainx.gph" "$results/ovr_sub_health_mainx.gph" , c(3) graphr(color(white)) xsize(4) ysize(2)  note("Individual FE-Regressions. Conditional on: Interview month, interview mode, age (linear).")

graph export "$results/other_wave.png", replace
graph export "$results/other_wave.pdf", replace
graph export "$results/other_wave.wmf", replace
```
