## Supplementary material for "Mental health in Germany before, during and after the COVID-19 pandemic": Replication Syntax: readme.docx

**Replication Syntax for the Paper “Mental health in Germany before, during and after the COVID-19 pandemic” published in PLOS One**

Authors: Alexander Patzina, Matthias Collischon, Rasmus Hoffmann, Maksym Obrizan

The survey data from the PASS can be obtained by the Research Data centre of the IAB at: <https://fdz.iab.de/pd_hd/panel-arbeitsmarkt-und-soziale-sicherung-pass-version-0622-v1/>

The syntax simply recreates the Figures and Tables from the paper. For the analysis to run properly, the *esttab-*ado file in Stata is required to creates tables. Additionally, the *coefplot*-ado file is required to recreate the Figures.
